# DNA Methylation-Derived Immune Cell Proportions and Cancer Risk, Including Lung Cancer, in Black Participants

**DOI:** 10.1101/2024.05.09.24307118

**Authors:** Christopher S. Semancik, Naisi Zhao, Devin C. Koestler, Eric Boerwinkle, Jan Bressler, Rachel J. Buchsbaum, Karl T. Kelsey, Elizabeth A. Platz, Dominique S. Michaud

**Affiliations:** Department of Public Health & Community Medicine, Tufts University School of Medicine, Tufts University, Boston, MA, USA; The University of Kansas Cancer Center, Kansas City, KS, USA; Department of Biostatistics & Data Science, University of Kansas Medical Center, Kansas City, KS, USA; Human Genetics Center, School of Public Health, University of Texas Health Science Center at Houston, Houston, TX, USA; Human Genome Sequencing Center, Baylor College of Medicine, Houston, TX, USA; Division of Hematology/Oncology, Tufts Medical Center, Boston, MA, USA; Department of Epidemiology, Brown University, Providence, RI, USA; Department of Pathology and Laboratory Medicine, Brown University, Providence, RI, USA; Department of Epidemiology, Johns Hopkins Bloomberg School of Public Health, Baltimore, MD, USA; The Sidney Kimmel Comprehensive Cancer Center at Johns Hopkins, Baltimore, MD, USA

**Keywords:** DNA methylation, immune cell profiles, cancer risk, immunology, epidemiology

## Abstract

Prior cohort studies assessing cancer risk based on immune cell subtype profiles have predominantly focused on White populations. This limitation obscures vital insights into how cancer risk varies across race. Immune cell subtype proportions were estimated using deconvolution based on leukocyte DNA methylation markers from blood samples collected at baseline on participants without cancer in the Atherosclerosis Risk in Communities (ARIC) Study. Over a mean of 17.5 years of follow-up, 668 incident cancers were diagnosed in 2,467 Black participants. Cox proportional hazards regression was used to examine immune cell subtype proportions and overall cancer incidence and site-specific incidence (lung, breast, and prostate cancers). Higher T regulatory cell proportions were associated with statistically significantly higher lung cancer risk (hazard ratio = 1.22, 95% confidence interval = 1.06-1.41 per percent increase). Increased memory B cell proportions were associated with significantly higher risk of prostate cancer (1.17, 1.04-1.33) and all cancers (1.13, 1.05-1.22). Increased CD8+ naïve cell proportions were associated with significantly lower risk of all cancers in participants ≥55 years (0.91, 0.83-0.98). Other immune cell subtypes did not display statistically significant associations with cancer risk. These results in Black participants align closely with prior findings in largely White populations. Findings from this study could help identify those at high cancer risk and outline risk stratifying to target patients for cancer screening, prevention, and other interventions. Further studies should assess these relationships in other cancer types, better elucidate the interplay of B cells in cancer risk, and identify biomarkers for personalized risk stratification.

## Introduction

The immune system plays a key role in protecting against cancer ^1^. Studies using animal models and cancer patients have shown the immune system’s ability to recognize and eliminate tumor cells through immunosurveillance, which involves both innate and adaptive immune response ^2–4^. However, tumor cells can evade immunosurveillance responses by suppressing the immune system ^5,6^. Within tumors, higher proportions of cytotoxic T lymphocytes (CD8+ cells) have been associated with more favorable cancer outcomes, whereas higher proportions of T regulatory cells (Tregs) have been associated with immunosuppression, accelerated cancer development, and decreased survival ^7–10^. Intertumoral accumulation of Tregs has been consistently associated with greater tumor aggressiveness in patients with various cancer types ^11–14^.

Despite these insights, the role of peripheral blood immune cell profiles in the pre-cancerous state and their influence on subsequent cancer risk remains unclear. Further, cohort studies cannot systematically use flow cytometry to identify immune cell type profiles in peripheral blood at regular, consistent intervals because cohorts seldom have whole blood stored under appropriate conditions. To date, few observational studies have examined the relationship between immune cells measured in pre-diagnostic blood and cancer risk.

Recent advances in high-dimensional arrays enable measurement of DNA methylation at 450,000 to 850,000 CpG oligodeoxynucleotide sites (CpGs) throughout the genome, allowing precise estimates of immune cell proportions from frozen blood samples ^15–18^. Pre-diagnostic blood collection is essential to assess DNA methylation states and immune cell proportions because the cancers themselves may alter these profiles ^19^. Located throughout the genome, differentially methylated regions (DMRs) can distinctly identify the lineage of differentiated immune cell subtypes ^20^. These unique differentially methylated CpGs can be used to identify immune cell lineages, and their proportions can be estimated using a statistical method called “deconvolution” ^20^. Resulting immune cell proportions can be employed to assess relationships between immune cell profiles and cancer risk ^20^. Since DNA methylation analysis can be done using DNA from archived blood, immune profiles can now be assessed using the resources of large epidemiologic studies that have banked specimens.

For this analysis, we utilized the Atherosclerosis Risk in Communities (ARIC) study to investigate the risk of lung cancer, breast cancer, prostate cancer, and all cancers pooled together (excluding hematological cancers) in relation to DNA methylation-derived relative proportions of peripheral blood immune cell types in Black participants. This represents an important opportunity to analyze this relationship in an understudied population, as Black individuals are known to have lower average neutrophil counts than White individuals ^21,22^.

## Materials and Methods

### Study Population

Participants were members of the ARIC study (RRID: SCR_021769), a prospective cohort study of cardiovascular disease risk that enrolled 15,792 people between 1987 and 1989 from four different communities in the United States (Jackson, MS; Washington County, MD; suburban Minneapolis, MN; and Forsyth County, NC) ^23,24^. Participants underwent a baseline clinical examination (Visit 1; 1987-1989), which included an in-home interview and clinical examination, assessing medical and lifestyle factors ^25^. Participants returned for follow-up clinical examinations in 1990-1992 (Visit 2), 1993-1995 (Visit 3), 1996-1998 (Visit 4), 2011-2013 (Visit 5), and so on until Visit 10. Blood specimens were banked at each visit, and participants were followed by annual telephone calls until 2011 with semi-annual contact thereafter. The ARIC study protocol was approved by institutional review boards at each site and participants gave informed consent.

For this analysis, we included Black participants (initial total of n = 2,520) from the Jackson, MS community (n = 2,287) and the Forsyth County, NC community (n = 233) who previously had methylation profiling performed, fully consented to cancer and genetic research, and had no cancer history before blood collection.

### DNA Methylation Profiling and Prediction of Immune Cell Proportions

At Visit 2 or Visit 3, DNA methylation levels were measured ^26,27^. Then, using lineage CpG markers for immune subsets, immune cell deconvolution was conducted to estimate proportions for 12 leukocyte subtypes: neutrophils, eosinophils, basophils, monocytes, memory and naïve B cells, CD4+ and CD8+ naïve and memory cells, natural killer cells, and Tregs ^26^. Deconvolution has been validated against flow cytometry ^15,26,28–30^. Implausibly low immune cell proportion values were assigned the limit of detection, as described by Bell-Glenn and colleagues ^31^. Values for the methylation-derived neutrophil-to-lymphocyte ratio (mdNLR) were calculated by dividing neutrophil proportions by lymphocyte proportions, and are represented as a ratio. The R function EstDimRMT was utilized to determine the number of principal components with non-zero eigenvalues needed to correct batch effects ^32^.

### Cancer Ascertainment

Based on cancer registry data and follow-up, cancer cases were ascertained for all cancer types, except for non-melanoma skin cancer. The outcome of interest, cancer incidence of any type, was positive if a participant developed any type of cancer over the follow-up period. The only outcome of interest was first incidences of cancer, not cancer recurrences. For some analyses, the outcome was restricted to the most common types of cancer, including lung cancer, breast cancer, and prostate cancers. In the case of breast cancer individually, cases in premenopausal women were excluded due to small numbers, meaning all included cases of breast cancer individually occurred in postmenopausal women. The premenopausal breast cancer cases, however, were included in all cancers combined. Besides lung, breast, and prostate cancers, other individual cancers were not analyzed due to the low numbers of cases associated with other types of cancer, thus resulting in low and reduced statistical power if these cancers were to be analyzed individually.

Incident cancers were ascertained from baseline (either Visit 2 or Visit 3) until the end of 2015 through linkage with state cancer registries in Minnesota, North Carolina, Maryland, and Mississippi. From baseline through 2015, we ascertained 721 primary cancer cases and 345 cancer deaths in Black participants. These cases occurred over a mean of 17.5 years of follow-up. For analyses, all hematological cancers (n = 53) were removed, as hematological cancers originate in the progenitors that give rise to immune cells and may result in spurious immune profiles. This left 2,467 participants and 668 cancers for analysis.

### Covariate Assessment

Risk factors associated with cancer include: age, sex, BMI, cigarette smoking (self-reported smoking status, self-reported pack-years, and methylation-derived pack-years), postmenopausal hormone use, and alcohol consumption (self-reported drinking status). Data on cigarette smoking and drinking status (current, former, never) and cigarette smoking cumulative dose (pack-years) were collected at each visit during follow-up, and we used the corresponding data from the visit of blood draw. Postmenopausal hormone use at Visit 2 was used.

### Estimation of Peripheral Blood Leukocyte Composition

Briefly, the Illumina HumanMethylation450 BeadChip array was used for genome-wide DNA methylation profiling in 2,853 Black participants at 483,525 CpG sites. Genomic DNA was extracted from peripheral blood leukocyte samples using the Gentra Puregene Blood Kit (Qiagen), and bisulfite conversion of 1 µg genomic DNA was performed using the EZ-96 DNA Methylation Kit (Deep Well Format) (Zymo Research) ^27^.

As the main exposures of interest, peripheral blood leukocyte subtype proportions were estimated, including myeloid lineage subtypes (neutrophils, eosinophils, basophils, and monocytes) and lymphoid lineage subtypes (B lymphocytes naïve, B lymphocytes memory, T helper lymphocytes naïve (CD4+ naïve cells), T helper lymphocytes memory (CD4+ memory cells), T regulatory cells (Tregs), T cytotoxic lymphocytes naïve (CD8+ naïve cells), T cytotoxic lymphocytes memory (CD8+ memory cells), and natural killer lymphocytes). This estimation was done using a newly expanded reference-based deconvolution library EPIC IDOL-Ext. ^26^. This library used the IDOL methodology to optimize the currently available six-cell reference library, to deconvolve the proportions of 12 leukocyte subtypes in peripheral blood ^33,34^. This EPIC IDOL-Ext library was validated using gold standard flow cytometry data and substantiated by including publicly available data from more than 100,000 samples ^26^.

### Measurement of Complete Blood Count and Total Leukocyte Count

In addition to using the deconvolution technique to estimate peripheral blood leukocyte composition for each participant, complete blood count (CBC) was measured on archived blood of participants from Visit 3. For most participants, CBC was measured without differential analysis; 30 participants did not have CBC (n = 2,437 participants with CBC data).

### Statistical Analysis

To estimate the association of methylation-derived immune cell proportions with cancer incidence, we used Cox proportional hazards regression to estimate hazard ratios (HRs) and 95% confidence intervals (CIs) of total cancer, lung cancer, breast cancer, and prostate cancer, adjusting for known risk factors described in the covariate assessment. Participants contributed time at risk from blood draw for profiling at Visit 2 (89.1% of participants) or Visit 3 (10.9% of participants) until cancer diagnosis of any site, death, or administrative censoring at the end of 2015, whichever came first.

Specifically, the covariates included: age (continuous), sex, BMI (continuous), cigarette smoking status, cigarette smoking dose (continuous), postmenopausal hormone use, methylation-derived pack-years (continuous), methylation-derived neutrophil-to-lymphocyte ratio (mdNLR) (continuous), drinking status (for breast cancer only), and batch effect (continuous).

Analyses were performed using R Project for Statistical Computing (v4.0.2; R Core Team 2020 [RRID: SCR_001905]). Statistical tests were two-sided, and a P-value of less than 0.05 was considered statistically significant.

## Results

In this population, being older, a smoker (current smoker or higher pack-years), not using menopausal hormones (among women), having less education, or having a lower mdNLR was more common with increasing proportion of Tregs (**Table 1**). Being younger, female, a menopausal hormone user (among women), having more education, or having a lower mdNLR was more common with increasing CD8+ naïve cell proportions (**Table 2**).

**Table 1:** Baseline Characteristics for the Study Population, Characterized by Treg Proportion, Black Participants in ARIC.

| Characteristic |  | Quartile of Treg Composition |  |  |  | Overall<br>(n = 2467) |
| --- | --- | --- | --- | --- | --- | --- |
|  |  | Quartile 1<br>(n = 845) | Quartile 2<br>(n = 391) | Quartile 3<br>(n = 614) | Quartile 4<br>(n = 617) |  |
| **Age** |  |  |  |  |  |  |
|  | Mean (SD) | 55.8 (5.74) | 56.3 (5.86) | 56.4 (5.80) | 57.4 (5.73) | 56.4 (5.80) |
|  | Median | 55.0 | 56.0 | 56.0 | 57.0 | 56.0 |
|  | [Min, Max] | [47.0, 71.0] | [47.0, 69.0] | [47.0, 69.0] | [47.0, 70.0] | [47.0, 71.0] |
| **BMI** |  |  |  |  |  |  |
|  | Mean (SD) | 30.0 (5.76) | 30.0 (6.36) | 30.6 (6.67) | 29.8 (6.31) | 30.1 (6.25) |
|  | Median | 29.2 | 28.9 | 29.5 | 29.2 | 29.2 |
|  | [Min, Max] | [17.7, 53.8] | [15.1, 62.4] | [16.1, 57.9] | [14.7, 54.8] | [14.7, 62.4] |
| **Sex (%)** |  |  |  |  |  |  |
|  | Female | 529 (62.6%) | 239 (61.1%) | 399 (65.0%) | 401 (65.0%) | 1568 (63.6%) |
|  | Male | 316 (37.4%) | 152 (38.9%) | 215 (35.0%) | 216 (35.0%) | 899 (36.4%) |
| **Smoking Status (%)** |  |  |  |  |  |  |
|  | Never | 487 (57.6%) | 183 (46.8%) | 291 (47.3%) | 265 (42.9%) | 1226 (49.7%) |
|  | Former smoker | 193 (22.8%) | 98 (25.1%) | 171 (27.9%) | 164 (26.6%) | 626 (25.4%) |
|  | Current smoker | 165 (19.6%) | 110 (28.1%) | 152 (24.8%) | 188 (30.5%) | 615 (24.9%) |
| **Smoking Pack-Years, Current and Former Smokers** |  |  |  |  |  |  |
|  | Mean (SD) | 21.0 (23.4) | 20.9 (21.6) | 19.78 (18.7) | 25.5 (25.2) | 21.9 (22.6) |
|  | Median | 14.6 | 15.7 | 16.0 | 19.9 | 16.3 |
|  | [Min, Max] | [0, 199] | [0, 180] | [0, 138] | [0, 189] | [0, 199] |
| **Methylation-Derived Pack-Years** |  |  |  |  |  |  |
|  | Mean (SD) | 19.1 (34.5) | 21.5 (34.9) | 19.3 (35.6) | 22.5 (37.1) | 20.4 (35.5) |
|  | Median | 0 | 0 | 0 | 0 | 0 |
|  | [Min, Max] | [0, 302] | [0, 183] | [0, 239] | [0, 202] | [0, 302] |
| **Menopausal Hormone Therapy Use, Women Only (%)** |  |  |  |  |  |  |
|  | Never | 305 (57.7%) | 142 (59.4%) | 233 (58.4%) | 274 (68.3%) | 954 (60.8%) |
|  | Former user | 17 (3.2%) | 8 (3.3%) | 13 (3.3%) | 6 (1.5%) | 44 (2.8%) |
|  | Current user | 118 (22.3%) | 47 (19.7%) | 78 (19.5%) | 48 (12.0%) | 291 (18.6%) |
|  | Unknown | 89 (16.8%) | 42 (17.6%) | 75 (18.8%) | 73 (18.2%) | 279 (17.8%) |
| **Education Level (%)** |  |  |  |  |  |  |
|  | Basic Education (less than completed high school) | 269 (31.8%) | 160 (40.9%) | 252 (41.0%) | 290 (47.0%) | 971 (39.4%) |
|  | Intermediate Education (high school or equivalent) | 245 (29.0%) | 109 (27.9%) | 182 (29.6%) | 168 (27.2%) | 704 (28.5%) |
|  | Advanced Education (at least some college) | 331 (39.2%) | 119 (30.4%) | 176 (28.7%) | 158 (25.6%) | 784 (31.8%) |
|  | Missing | 0 (0%) | 3 (0.8%) | 4 (0.7%) | 1 (0.2%) | 8 (0.3%) |
| **Drinking Status (%)** |  |  |  |  |  |  |
|  | Never | 316 (37.4%) | 134 (34.3%) | 217 (35.3%) | 200 (32.4%) | 867 (35.1%) |
|  | Former drinker | 244 (28.9%) | 124 (31.7%) | 202 (32.9%) | 211 (34.2%) | 781 (31.7%) |
|  | Current drinker | 285 (33.7%) | 133 (34.0%) | 195 (31.8%) | 206 (33.4%) | 819 (33.2%) |
| **Methylation-Derived Neutrophil-to-Lymphocyte Ratio** |  |  |  |  |  |  |
|  | Mean (SD) | 3.22 (6.43) | 1.99 (2.24) | 1.67 (1.31) | 1.46 (0.967) | 2.21 (4.02) |
|  | Median | 1.36 | 1.37 | 1.34 | 1.17 | 1.32 |
|  | [Min, Max] | [0.223, 46.3] | [0.308, 33.1] | [0.176, 12.2] | [0.263, 11.5] | [0.176, 46.3] |

**Table 2:** Baseline Characteristics for the Study Population, Characterized by CD8+ Naïve Cell Proportion, Black Participants in ARIC.

| Characteristic |  | Quartile of CD8+ Naïve Cell Composition |  |  |  | Overall<br>(n = 2467) |
| --- | --- | --- | --- | --- | --- | --- |
|  |  | Quartile 1<br>(n = 1111) | Quartile 2<br>(n = 452) | Quartile 3<br>(n = 452) | Quartile 4<br>(n = 452) |  |
| **Age** |  |  |  |  |  |  |
|  | Mean (SD) | 57.7 (5.91) | 56.2 (5.62) | 55.6 (5.44) | 54.3 (5.28) | 56.4 (5.80) |
|  | Median | 58.0 | 56.0 | 55.0 | 53.0 | 56.0 |
|  | [Min, Max] | [47.0, 71.0] | [47.0, 70.0] | [47.0, 70.0] | [47.0, 69.0] | [47.0, 71.0] |
| **BMI** |  |  |  |  |  |  |
|  | Mean (SD) | 30.2 (6.28) | 30.2 (6.67) | 29.7 (5.84) | 30.2 (6.03) | 30.1 (6.23) |
|  | Median | 29.3 | 29.4 | 28.9 | 29.2 | 29.3 |
|  | [Min, Max] | [14.7, 54.9] | [15.1, 62.4] | [15.8, 53.8] | [16.1, 52.8] | [14.7, 62.4] |
| **Sex (%)** |  |  |  |  |  |  |
|  | Female | 638 (57.4%) | 267 (59.1%) | 321 (71.0%) | 342 (75.7%) | 1568 (63.6%) |
|  | Male | 473 (42.6%) | 185 (40.9%) | 131 (29.0%) | 110 (24.3%) | 899 (36.4%) |
| **Smoking Status (%)** |  |  |  |  |  |  |
|  | Never | 572 (51.5%) | 219 (48.6%) | 222 (49.1%) | 213 (47.1%) | 1226 (49.7%) |
|  | Former smoker | 277 (24.9%) | 116 (25.7%) | 117 (25.9%) | 116 (25.7%) | 626 (25.4%) |
|  | Current smoker | 262 (23.6%) | 117 (25.9%) | 113 (25.0%) | 123 (27.2%) | 615 (24.9%) |
| **Smoking Pack-Years, Current and Former Smokers** |  |  |  |  |  |  |
|  | Mean (SD) | 23.1 (22.6) | 22.3 (22.6) | 22.0 (26.8) | 18.9 (17.5) | 21.9 (22.6) |
|  | Median | 18.6 | 16.9 | 15.2 | 14.0 | 16.3 |
|  | [Min, Max] | [0,199] | [0, 160] | [0, 189] | [0, 82.0] | [0, 199] |
| **Methylation-Derived Pack-Years** |  |  |  |  |  |  |
|  | Mean (SD) | 20.8 (36.0) | 19.8 (35.1) | 18.8 (33.7) | 21.5 (36.6) | 20.4 (35.5) |
|  | Median | 0 | 0 | 0 | 0 | 0 |
|  | [Min, Max] | [0, 302] | [0, 188] | [0, 167] | [0, 239] | [0, 302] |
| **Menopausal Hormone Therapy Use, Women Only (%)** |  |  |  |  |  |  |
|  | Never | 401 (62.8%) | 162 (60.7%) | 188 (58.6%) | 203 (59.4%) | 954 (60.8%) |
|  | Former user | 12 (1.9%) | 10 (3.7%) | 10 (3.1%) | 12 (3.5%) | 44 (2.8%) |
|  | Current user | 111 (17.4%) | 50 (18.7%) | 57 (17.8%) | 73 (21.3%) | 291 (18.6%) |
|  | Unknown | 114 (17.9%) | 45 (16.9%) | 66 (20.5%) | 54 (15.8%) | 279 (17.8%) |
| **Education Level (%)** |  |  |  |  |  |  |
|  | Basic Education (less than completed high school) | 492 (44.3%) | 168 (37.2%) | 179 (39.6%) | 132 (29.2%) | 971 (39.4%) |
|  | Intermediate Education (high school or equivalent) | 304 (27.4%) | 137 (30.3%) | 135 (29.9%) | 128 (28.3%) | 704 (28.5%) |
|  | Advanced Education (at least some college) | 309 (27.8%) | 146 (32.3%) | 138 (30.5%) | 191 (42.3%) | 784 (31.8%) |
|  | Missing | 6 (0.5%) | 1 (0.2%) | 0 (0%) | 1 (0.2%) | 8 (0.3%) |
| **Drinking Status (%)** |  |  |  |  |  |  |
|  | Never | 366 (32.9%) | 153 (33.8%) | 170 (37.6%) | 178 (39.4%) | 867 (35.1%) |
|  | Former drinker | 376 (33.8%) | 153 (37.6%) | 135 (29.9%) | 118 (26.1%) | 781 (31.7%) |
|  | Current drinker | 369 (33.3%) | 147 (32.5%) | 147 (32.5%) | 156 (34.5%) | 819 (33.2%) |
| **Methylation-Derived Neutrophil-to-Lymphocyte Ratio** |  |  |  |  |  |  |
|  | Mean (SD) | 2.61 (5.27) | 2.44 (3.58) | 1.91 (2.25) | 1.38 (1.11) | 2.21 (4.02) |
|  | Median | 1.42 | 1.51 | 1.30 | 0.984 | 1.32 |
|  | [Min, Max] | [0.263, 46.3] | [0.176, 37.5] | [0.230, 26.2] | [0.223, 9.92] | [0.176, 46.3] |

Looking at cancer risks overall based on all 12 immune cell subtypes, many findings were not statistically significant (**Table 3**). However, three cell subtypes especially demonstrated statistically significant results when analyzed as continuous variables: Tregs, CD8+ naïve cells, and memory B cells. Further, total leukocyte counts (derived from complete blood count) exhibited positive statistical significance.

**Table 3:** Hazard Ratios (HRs) for Immune Cell Proportions and Cancer Risk, Black Participants in ARIC. HR ^b^ (95% CI) per 1 percent increase in methylation-derived immune cell proportion or 1 unit increase in ratios or white blood cell count

| Methylation-Derived Immune Cell Type or Measure | All Cancer<sup>a</sup><br>(668 cases) | Lung Cancer<br>(84 cases) | Postmenopausal Breast Cancer<sup>c</sup><br>(114 cases) | Prostate Cancer<sup>d</sup><br>(173 cases) |
| --- | --- | --- | --- | --- |
| CD4+ | 0.99 (0.98, 1.01) | 1.00 (0.96, 1.03) | 1.00 (0.97, 1.02) | 1.00 (0.98, 1.03) |
| CD4+ memory | 0.99 (0.98, 1.01) | 0.99 (0.95, 1.04) | 1.01 (0.98, 1.05) | 1.00 (0.97, 1.03) |
| CD4+ naïve | 1.00 (0.98, 1.02) | 1.00 (0.95, 1.06) | 0.97 (0.93, 1.01) | 1.01 (0.97, 1.06) |
| CD4+ naïve-to-memory ratio | 1.02 (0.98, 1.05) | 0.96 (0.85, 1.09) | 1.00 (0.93, 1.08) | 0.98 (0.90, 1.06) |
| CD8+ | 1.00 (0.99, 1.01) | 1.02 (0.99, 1.06) | 1.02 (0.98, 1.05) | 1.01 (0.99, 1.03) |
| CD8+ memory | 1.00 (0.99, 1.02) | 1.03 (1.00, 1.06) | 1.02 (0.99, 1.05) | 1.01 (0.99, 1.03) |
| CD8+ naïve | 0.96 (0.91, 1.01) | 0.85 (0.71, 1.01) | 0.96 (0.86, 1.07) | 1.00 (0.89, 1.12) |
| CD8+ naïve-to-memory ratio | 0.98 (0.87, 1.09) | 0.87 (0.59, 1.29) | 0.96 (0.75, 1.24) | 0.84 (0.62, 1.13) |
| CD4+-to-CD8+ ratio | 0.97 (0.91, 1.04) | 0.98 (0.84, 1.15) | 0.89 (0.75, 1.07) | 0.94 (0.82, 1.07) |
| Treg | 1.06 (1.00, 1.12) | **1.22 (1.06, 1.41)** | 1.04 (0.92, 1.18) | 1.08 (0.96, 1.22) |
| B cell | 0.99 (0.97, 1.02) | 0.98 (0.91, 1.06) | 1.04 (0.98, 1.10) | 0.97 (0.92, 1.03) |
| B cell memory | **1.13 (1.05, 1.22)** | 1.20 (0.99, 1.46) | 0.95 (0.74, 1.24) | **1.17 (1.04, 1.33)** |
| B cell naïve | 0.98 (0.95, 1.01) | 0.96 (0.88, 1.04) | 1.04 (0.98, 1.10) | 0.94 (0.89, 1.00) |
| B cell naïve-to-memory ratio | 0.99 (0.98, 1.00) | 1.06 (0.94, 1.00) | 1.01 (0.99, 1.04) | 0.98 (0.96, 1.00) |
| NLR | 1.00 (0.98, 1.02) | 0.95 (0.87, 1.03) | 0.99 (0.94, 1.04) | 1.01 (0.98, 1.04) |
| Lymphocyte to Monocyte Ratio | 1.00 (1.00, 1.00) | 1.00 (1.00, 1.00) | 1.00 (1.00, 1.00) | 1.03 (0.95, 1.12) |
| White blood cell count (from CBC differential) | 1.03 (0.98, 1.08)<br>(n = 659) | **1.19 (1.06, 1.33)**<br>(n = 83) | 1.05 (0.94, 1.18)<br>(n = 112) | 0.99 (0.91, 1.08)<br>(n = 171) |

<sup>a</sup> Excluding hematologic cancers. <sup>b</sup> Multivariable models were adjusted for age, sex, BMI, self-reported smoking status, self-reported smoking pack-years, postmenopausal hormone use (creating an 'NA' category for males), methylation-derived smoking pack-years, mdNLR (in all models except mdNLR and lymphocyte to monocyte ratio), and batch effect. <sup>c</sup> The breast cancer model also adjusted for self-reported drinking status and did not adjust for sex. <sup>d</sup> The prostate cancer model did not adjust for sex or postmenopausal hormone use (men only).

Assessing Treg proportions, each one-percent increase was associated with a 6% increased risk of all cancers (HR: 1.06, 95% CI = 1.00 to 1.11) (**Table 3**). For lung cancer, a one-percent increase of Treg proportion was associated with a 22% increased risk of lung cancer (HR: 1.22, 95% CI = 1.06 to 1.41) (**Table 3**). Treg proportions were associated with increased risk of breast cancer and prostate cancer but were not statistically significant (**Table 3**). In a sensitivity analysis, we further adjusted for education level and diabetes status at baseline, and the associations in all cancers and lung cancer were not appreciably changed (**Supplementary Table 1**). Treg associations did not vary substantially when stratified by age (55 years old or younger versus more than 55 years old) or by sex (**Tables 4-6**; **Supplementary Tables 7-9**). Mutually adjusting for measured total leukocyte count and methylation-derived Treg proportions did not attenuate findings for either measure (**Supplementary Table 6**).

**Table 4:**
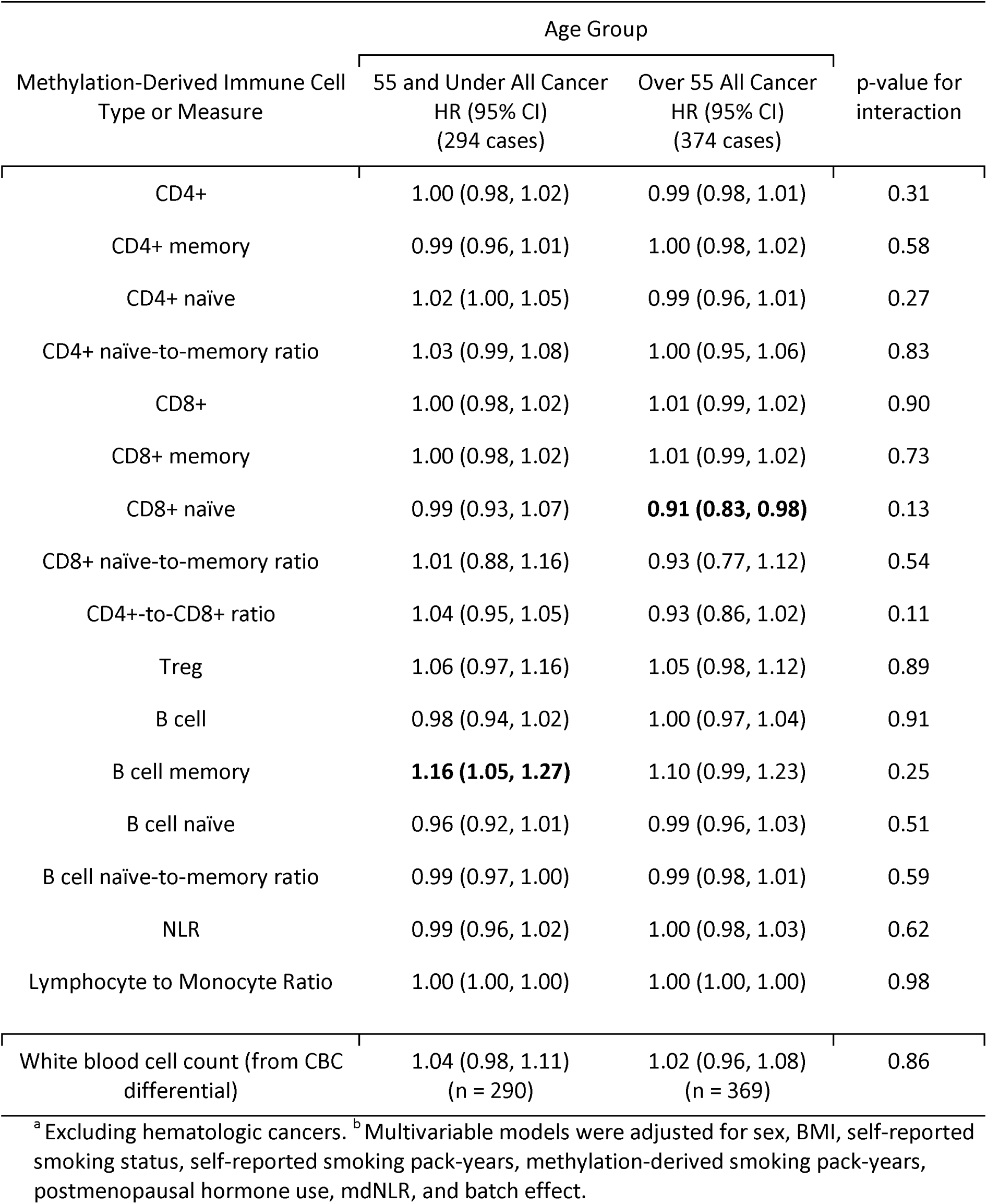
HRs for Age-Stratified Model in All Cancers ^a^, Black Participants in ARIC. HR ^b^ (95% CI) per 1 percent increase in methylation-derived immune cell proportion or 1 unit increase in ratios or white blood cell count

Analyzing CD8+ naïve cell proportions, a one-percent increase was associated with a 4% decreased risk of all cancers (HR: 0.96, 95% CI = 0.91 to 1.01) (**Table 3**). Each one-percent increase in CD8+ naïve cell proportion was associated with a 15% decreased risk of lung cancer (HR: 0.85, 95% CI = 0.71 to 1.01; **Table 3**). CD8+ naïve cell proportion was not associated with breast cancer or prostate cancer risk (**Table 3**). In a sensitivity analysis, we further adjusted for education level and diabetes status at baseline, and associations in all cancers and lung cancer were not appreciably changed (**Supplementary Table 1**). However, for age-stratified analyses, results were statistically significant in all cancers for individuals over 55 years old (HR: 0.91, 95% CI = 0.83 to 0.98), but null for individuals 55 years old or younger, and cases were similarly distributed by age group (**Table 4**). Age-stratified results were directionally similar in individuals over 55 years old, but not statistically significant, in the individual cancers assessed (**Tables 5-6**).

**Table 5:** HRs for Age-Stratified Model in Postmenopausal Breast Cancer, Black Participants in ARIC. HR ^a^ (95% CI) per 1 percent increase in methylation-derived immune cell proportion or 1 unit increase in ratios or white blood cell count

| Methylation-Derived Immune Cell Type or Measure | Age Group |  | p-value for interaction |
| --- | --- | --- | --- |
|  | 55 and Under Breast Cancer HR (95% CI) (49 cases) | Over 55 Breast Cancer HR (95% CI) (65 cases) |  |
| CD4+ | 0.99 (0.94, 1.04) | 1.00 (0.96, 1.03) | 0.67 |
| CD4+ memory | 1.00 (0.95, 1.06) | 1.02 (0.97, 1.06) | 0.65 |
| CD4+ naïve | 0.98 (0.92, 1.05) | 0.96 (0.90, 1.02) | 0.76 |
| CD4+ naïve-to-memory ratio | 1.04 (0.95, 1.13) | 0.95 (0.83, 1.09) | 0.64 |
| CD8+ | 1.03 (0.98, 1.08) | 1.01 (0.97, 1.05) | 0.19 |
| CD8+ memory | 1.02 (0.98, 1.07) | 1.01 (0.98, 1.05) | 0.23 |
| CD8+ naïve | 1.00 (0.85, 1.17) | 0.90 (0.75, 1.07) | 0.68 |
| CD8+ naïve-to-memory ratio | 1.09 (0.82, 1.45) | 0.79 (0.48, 1.28) | 0.40 |
| CD4+-to-CD8+ ratio | 0.91 (0.68, 1.22) | 0.89 (0.71, 1.11) | 0.83 |
| Treg | 1.09 (0.90, 1.32) | 1.02 (0.87, 1.19) | 0.60 |
| B cell | 0.98 (0.89, 1.09) | 1.06 (0.99, 1.14) | 0.93 |
| B cell memory | 0.92 (0.64, 1.30) | 0.95 (0.64, 1.42) | 0.54 |
| B cell naïve | 0.99 (0.90, 1.10) | 1.07 (1.00, 1.15) | 0.82 |
| B cell naïve-to-memory ratio | 1.00 (0.96, 1.03) | 1.03 (1.00, 1.05) | 0.46 |
| NLR | 0.88 (0.71, 1.09) | 1.02 (0.97, 1.07) | 0.37 |
| Lymphocyte to Monocyte Ratio | 1.00 (1.00, 1.00) | 1.00 (1.00, 1.00) | 0.67 |
| White blood cell count (from CBC differential) | 1.09 (0.92, 1.30) (n = 48) | 1.02 (0.87, 1.20) (n = 64) | 0.85 |

<sup>a</sup> Multivariable models were adjusted for BMI, self-reported smoking status, self-reported smoking pack-years, methylation-derived smoking pack-years, self-reported drinking status, postmenopausal hormone use, mdNLR, and batch effect.

**Table 6:** HRs for Age-Stratified Model in Prostate Cancer, Black Participants in ARIC. HR ^a^ (95% CI) per 1 percent increase in methylation-derived immune cell proportion or 1 unit increase in ratios or white blood cell count

| Methylation-Derived Immune Cell Type or Measure | Age Group |  | p-value for interaction |
| --- | --- | --- | --- |
|  | 55 and Under Prostate Cancer HR (95% CI) (79 cases) | Over 55 Prostate Cancer HR (95% CI) (94 cases) |  |
| CD4+ | 1.01 (0.97, 1.05) | 1.00 (0.97, 1.03) | 0.06 |
| CD4+ memory | 0.99 (0.94, 1.03) | 1.01 (0.97, 1.05) | 0.26 |
| CD4+ naïve | 1.05 (0.98, 1.12) | 0.99 (0.93, 1.05) | 0.12 |
| CD4+ naïve-to-memory ratio | 1.00 (0.88, 1.14) | 0.96 (0.85, 1.08) | 0.83 |
| CD8+ | 1.00 (0.97, 1.01) | 1.01 (0.98, 1.04) | 0.38 |
| CD8+ memory | 1.00 (0.96, 1.03) | 1.01 (0.99, 1.04) | 0.68 |
| CD8+ naïve | 1.07 (0.95, 1.01) | 0.81 (0.64, 1.04) | 0.02 |
| CD8+ naïve-to-memory ratio | 0.94 (0.68, 1.29) | 0.61 (0.32, 1.17) | 0.28 |
| CD4+-to-CD8+ ratio | 1.02 (0.86, 1.20) | 0.86 (0.71, 1.06) | 0.22 |
| Treg | 1.08 (0.89, 1.09) | 1.07 (0.91, 1.25) | 0.37 |
| B cell | 1.00 (0.92, 1.09) | 0.94 (0.87, 1.02) | 0.04 |
| B cell memory | **1.25 (1.10, 1.43)** | 0.98 (0.77, 1.26) | 0.09 |
| B cell naïve | 0.94 (0.85, 1.03) | 0.94 (0.86, 1.02) | 0.36 |
| B cell naïve-to-memory ratio | 0.97 (0.94, 1.01) | 0.99 (0.96, 1.02) | 0.88 |
| NLR | 0.96 (0.88, 1.05) | 1.03 (0.99, 1.06) | 0.10 |
| Lymphocyte to Monocyte Ratio | 1.11 (0.94, 1.31) | 1.00 (0.99, 1.01) | 0.04 |
| White blood cell count (from CBC differential) | 1.01 (0.88, 1.15) (n = 77) | 0.98 (0.87, 1.10) (n = 94) | 0.22 |

<sup>a</sup> Multivariable models represent hazard ratios for each one-percent increase in cell subtype composition in prostate cancer; models were adjusted for BMI, self-reported smoking status, self-reported smoking pack-years, methylation-derived smoking pack-years, mdNLR, and batch effect.

Assessing memory B cell proportions, each one-percent increase was associated with a 13% increased risk of all cancers (HR: 1.13, 95% CI = 1.05 to 1.22), a 17% increased risk of prostate cancer (HR: 1.17, 95% CI = 1.04 to 1.33), and a 20% increased risk of lung cancer (HR: 1.20, 95% CI = 0.99 to 1.46) (**Table 3**). Since prostate cancer affects some men minimally and others quite aggressively, an analysis of only prostate cancer cases without a lethal phenotype exhibited a similarly statistically significant memory B cell association (HR: 1.19, 95% CI = 1.05, 1.34). Increasing memory B cell proportion was suggestive of reduced breast cancer risk, although this relationship was not statistically significant. Associations were similar in all cancers, prostate cancer, and lung cancer after further adjusting for education level and diabetes status (**Supplementary Table 1**). However, for age-stratified analyses, results were statistically significant in all cancers (HR: 1.16, 95% CI = 1.05 to 1.27) and prostate cancer (HR: 1.25, 95% CI = 1.10 to 1.43) for individuals 55 years old or younger, but null for individuals over 55 years old (**Tables 4**, **6**). Age-stratified results were not statistically significant in breast cancer, although the relationships were suggestively protective in both age groups (**Table 5**). The associations in men were statistically significant for all cancers (HR: 1.16, 95% CI = 1.05 to 1.28) but not women, and in lung cancer for men (HR: 1.34, 95% CI = 1.04 to 1.71) but not women; this relationship may be partly influenced by the direct contribution of prostate cancer cases in men only (**Supplementary Tables 7-8**).

Although the associations for Tregs, CD8+ naïve cells, and memory B cells for all cancers and lung cancer followed a linear dose-response (**Supplementary** Figures 1-2), spline plots showed that Treg and CD8+ naïve cell associations for breast and prostate cancers follow a non-linear pattern (**Supplementary** Figures 3-4). Therefore, we conducted quantile-based analyses for all cancers and individual cancers to address nuance in relationships modeled in previously discussed continuous models. In quartile analysis for all cancers, a 31% elevated risk of all cancers was observed for the highest quartile of Treg proportion, relative to the lowest quartile (HR: 1.31, 95% CI = 1.04 to 1.65) (**Supplementary Table 3**). Furthermore, a 47% increased risk of all cancers was observed for the highest compared with the lowest quartile of memory B cell proportion (HR: 1.47, 95% CI = 1.13 to 1.91) (**Supplementary Table 3**).

Using tertiles for the lung cancer analysis (due to a smaller number of cases), there was a dose-response, but not statistically significant relationship for Treg and memory B cell proportion, while for CD8+ naïve cell proportion, a statistically significant relationship was observed for the highest compared with the lowest tertile (HR: 0.47, 95% CI = 0.25 to 0.87) (**Supplementary Table 4**). For white blood cell count, a 167% elevated risk of lung cancer was observed for the highest compared with the lowest tertile of white blood cell count (HR: 2.67, 95% CI = 1.43 to 4.97) (**Supplementary Table 4**). For breast cancer, relationships were generally less dose-response oriented (**Supplementary Table 5**). Only white blood cell count followed a somewhat dose-response oriented relationship for breast cancer. For prostate cancer, associations were similarly non-linear (**Supplementary Table 6**).

## Discussion

Our study uniquely highlights that in a Black population, higher methylation-derived peripheral blood Treg proportion was associated with elevated risk of lung cancer, even when adjusting for smoking status and pack-years. Furthermore, for Treg proportion, a non-significant elevated risk of prostate cancer was noted. Additionally, increased memory B cell proportion was associated with elevated risk of all cancers and prostate cancer. Conversely, CD8+ naïve cell proportion was associated with a decreased risk of developing lung cancer and all cancers, an association that was statistically significant for individuals over 55 years old, suggesting that adaptive immunity in older age could be critical to combatting primary tumorigenesis. A positive association between total white blood cell count (directly measured) and lung cancer risk was also observed, which was independent of relationships measured in methylation-derived immune cell subtypes. Our findings extend prior findings which examined immune cell profiles and risk of major cancer types to a new population, using a cutting-edge algorithm for predicting immune cell subtype proportions.

Our work is among the first to investigate these associations in a Black cohort. Nonetheless, our findings merit comparison to other existing studies. Several studies have addressed the association between total white blood cell count and cancer risk. Studies conducted in the Women’s Health Initiative (WHI) and UK Biobank cohorts have reported that elevated white blood cell counts are associated with statistically significant increases in risk of invasive breast cancer in postmenopausal women, as well as endometrial cancer and lung cancer in a general cohort of individuals between 40 and 69 years old ^35,36^. Fewer studies have examined immune cell subtypes. Generally, higher proportions of Tregs relative to total leukocytes or other immune cells have been associated with higher risks of lung, colorectal, breast, and pancreatic cancers ^16–18^. On the other hand, higher relative proportions of CD8+ cells have been inversely associated with risk of lung cancer and breast cancer ^16,18^. Using a different cohort (the CLUE study), we previously detected a statistically significant increase in risk of non-small cell lung cancer for an increase of one standard deviation in mdNLR ^37^. Additionally, a 2020 paper by Kresovich et al. reported that increased B cell proportions are associated with higher breast cancer risk, and that increased monocyte proportions are associated with lower breast cancer risk among premenopausal women ^38^. In contrast, we did not observe any statistically significant relationships between four measures of immune cell proportions (mdNLR, CD4+/CD8+ total cells, B cells/lymphocytes, and T cells/lymphocytes) and risk of pancreatic cancer in a separate study ^19^.

In addition to observational studies, investigations into cancer cells, hosts, and microenvironments have postulated mechanisms through which cancerous cells are eliminated efficiently by CD8+ cells, while Tregs weaken cellular immune response by impeding the activation of T effector cells, preventing cancer cells from being destroyed and promoting tumor growth ^2,3,39^. This induces cellular and molecular networks, which induce an immunosuppressive environment that favors tumor growth ^40–42^. Increased ratios of Tregs to CD8+ cells have been shown to be an indicator of this immune evasion and tumor growth within the tumor microenvironment (TME) ^43–45^. It is still largely unknown how Tregs and B cells may interplay to initiate or accelerate tumorigenesis. On the other hand, CD8+ naïve cells are preferential immune cell types for targeting and preventing carcinogenesis ^46^. During carcinogenesis, CD8+ cells encounter dysfunction and exhaustion due to immune-related tolerance and immunosuppression within the TME, which favors adaptive immune resistance ^46^. Upon their activation, CD8+ cells infiltrate to the core of the tumor’s invading site and kill cancer cells ^46^. By killing malignant cells upon recognition of specific antigenic peptides by T-cell receptors, CD8+ cells play a central protective role in cancer immunity, unlike Tregs ^47^.

There are many key strengths of this analysis. First, the ARIC study is a prospective cohort with a large number of Black participants ^23^. Prior studies utilizing cohorts such as EPIC, WHI, UK Biobank, the Sister Study, and CLUE II have focused on White populations ^16–18,48^. To our knowledge, no existing study has evaluated the association between methylation-derived immune cell subtypes and cancer risk in a Black study population, extending prior findings into a historically under-researched population. Furthermore, many prior studies have focused only on major immune cell types, and did not study immune cell subtypes such as Tregs, CD8+ naïve cells, and memory B cells. This analysis utilizes the innovative deconvolution algorithm to predict proportions of 12 distinct immune cell subtypes ^19,39,46^. Unlike previously used methods, deconvolution allows for immune cell proportions to be estimated and measured from archived blood (peripheral blood leukocytes), allowing for a prospective study design that examines the role of systemic immune response in cancer risk ^18,20^.

One key limitation of this study is the relatively small number of individual cancer cases, such as lung cancer (n = 84 cases). Despite this, statistically significant associations observed indicate robust findings. However, larger sample sizes are necessary for a more detailed analysis of specific cancer types, such as colorectal cancer (n = 67 cases). We cannot rule out measurement error (including batch-to-batch variation in array data), residual confounding, and reverse causation (including undetected cancer incidence increasing immune cell proportions during short follow-up periods) ^18,20^. While cohort studies inherently carry risks of such biases, we have accounted for them through comprehensive multivariable models, sensitivity analyses (**Supplementary Tables 1, 7**), time-lag analyses (**Supplementary Table 2**), adjustment for batch effect by accounting for heterogeneity of measurements at CpG sites, and a biological measure of smoking pack-years. Taken together, accounting for these biases allows us to conclude that undetected, developing, or incident cancers were likely not driving increased or decreased cell proportions and that residual confounding or measurement error minimally contributed to the observed results. Using the Bonferroni correction for multiple comparisons (16 tests; adjusted significance level of P = 0.05/16 = 0.003), some findings are no longer statistically significant (Tregs in lung cancer and B memory cells in prostate cancer), so these results should be interpreted with caution as they could represent chance findings. However, our findings are largely consistent with prior studies, so results are likely not due to chance alone. Finally, because only postmenopausal women were included in breast cancer analysis, we could not directly replicate prior B cell findings in premenopausal women ^38^.

There are multiple areas for future study, chief among which is the need to identify biomarkers for personalized cancer risk stratification. This will permit better individualized decisions regarding preventive interventions, such as designing guidelines, laying out early screening regimens, and informing patients on lifestyle choices. Future research should strive to validate and refine deconvolution in Black individuals. Further studies should examine the impact of B cells on premenopausal breast cancer risk. Finally, larger studies in Black populations could confirm our findings and allow for assessment of other individual cancers.

In summary, this study shows that in a Black population, higher methylation-derived proportion of Tregs is associated with increased risk of lung cancer, higher memory B cell proportion is associated with increased risk of all cancers and prostate cancer, and higher CD8+ naïve cell proportion is associated with decreased risk of lung cancer and all cancers, particularly at older age. Our study underscores the complex interplay between various immune cell types and cancer risk in a Black population. These insights contribute significantly to our understanding of cancer immunology and highlight the need for further research in diverse populations to enhance cancer prevention and treatment strategies.

## Data Availability

Due to its proprietary nature, supporting data from the Atherosclerosis Risk in Communities cohort cannot be made openly available. Further information about the data and conditions for access are available at the Atherosclerosis Risk in Communities website at https://aric.cscc.unc.edu/aric9/.

## Acknowledgements

The authors thank the staff and participants of the ARIC study for their important contributions. Cancer data were provided by the Maryland Cancer Registry, Center for Cancer Prevention and Control, Maryland Department of Health, with funding from the State of Maryland and the Maryland Cigarette Restitution Fund. The collection and availability of cancer registry data is also supported by the Cooperative Agreement NU58DP007114, funded by the Centers for Disease Control and Prevention. Its contents are solely the responsibility of the authors and do not necessarily represent the official views of the Centers for Disease Control and Prevention or the Department of Health and Human Services.

## Author Contributions

The CRediT contribution roles that each author played in the development of this manuscript are detailed next to their initials below.

CSS: Data curation; formal analysis; investigation; methodology; validation; visualization; writing – original draft; writing – review & editing

NZ: Data curation; methodology; supervision; writing – review & editing

DCK: Software; supervision; writing – review & editing

EB: Funding acquisition; resources; writing – review & editing

JB: Writing – review & editing

RJB: Funding acquisition; writing – review & editing

KTK: Conceptualization; methodology; writing – review & editing

EAP: Conceptualization; funding acquisition; methodology; project administration; resources; supervision; writing – review & editing

DSM: Conceptualization; funding acquisition; investigation; methodology; project administration; supervision; validation; visualization; writing – original draft; writing – review & editing

The work reported in the paper has been performed by the authors, unless clearly specified in the text.

## List of Abbreviations (alphabetical order)

95% CI: 95% confidence interval
ARIC: Atherosclerosis Risk in Communities (cohort study)
CD8+ cells: cytotoxic T lymphocytes
CpG: CpG oligodeoxynucleotide sites
DMR: differentially methylated region
DNA: deoxyribonucleic acid
HR: hazard ratio
MD: Maryland
mdNLR: methylation-derived neutrophil-to-lymphocyte ratio
MN: Minnesota
MS: Mississippi
NC: North Carolina
SD: standard deviation
TME: tumor microenvironment
Tregs: T regulatory cells
WHI: Women’s Health Initiative (cohort study)

