## Supplemental Material for "DNA Methylation-Derived Immune Cell Proportions and Cancer Risk, Including Lung Cancer, in Black Participants"

**5/9/2024**

**Supplementary Materials for Article: DNA Methylation-Derived Immune Cell Proportions and Cancer Risk, Including Lung Cancer, in Black Participants**

Authors: Christopher S. Semancik, M.P.H., B.S.^1^, Naisi Zhao, M.S., Dr.PH^1^, Devin C. Koestler, Ph.D.^2,3^, Eric Boerwinkle, Ph.D.^4,5^, Jan Bressler, Ph.D.^4^, Rachel J. Buchsbaum, M.D.^6^, Karl T. Kelsey, M.D., M.O.H.^7,8^, Elizabeth A. Platz, Sc.D, M.P.H.^9,10^, Dominique S. Michaud, Sc.D.^1^

**Author Affiliations:**

^1^Department of Public Health & Community Medicine, Tufts University School of Medicine, Tufts University, Boston, MA, USA

^2^The University of Kansas Cancer Center, Kansas City, KS, USA

^3^Department of Biostatistics & Data Science, University of Kansas Medical Center, Kansas City, KS, USA

^4^Human Genetics Center, School of Public Health, University of Texas Health Science Center at Houston, Houston, TX, USA

^5^Human Genome Sequencing Center, Baylor College of Medicine, Houston, TX, USA

^6^Division of Hematology/Oncology, Tufts Medical Center, Boston, MA, USA

^7^Department of Epidemiology, Brown University, Providence, RI, USA

^8^Department of Pathology and Laboratory Medicine, Brown University, Providence, RI, USA

^9^Department of Epidemiology, Johns Hopkins Bloomberg School of Public Health, Baltimore, MD, USA

^10^The Sidney Kimmel Comprehensive Cancer Center at Johns Hopkins, Baltimore, MD, USA

**Corresponding Author:**

Dominique S. Michaud, ScD

Tufts University School of Medicine

136 Harrison Avenue

Boston, MA 02111

**Keywords:** DNA methylation, immune cell profiles, deconvolution, immunology, epidemiology

Supplementary Table 1: HRs for Sensitivity Analysis Including Diabetes and Educational Statuses:

HR ^b^ (95% CI) per 1 percent increase in methylation-derived immune cell proportion or 1 unit increase in ratios or white blood cell count

| Methylation-Derived Immune Cell Type or Measure | All Cancer ^a^  (668 cases) | Lung Cancer  (84 cases) | Postmenopausal Breast Cancer ^c^  (114 cases) | Prostate Cancer ^d^  (173 cases) |
| --- | --- | --- | --- | --- |
| CD4+ | 0.99 (0.98, 1.01) | 0.99 (0.96, 1.03) | 1.00 (0.97, 1.02) | 1.00 (0.98, 1.03) |
| CD4+ memory | 0.99 (0.98, 1.01) | 0.99 (0.95, 1.03) | 1.01 (0.98, 1.05) | 1.00 (0.97, 1.03) |
| CD4+ naïve | 1.00 (0.98, 1.02) | 1.00 (0.95, 1.06) | 0.97 (0.93, 1.02) | 1.01 (0.96, 1.05) |
| CD4+ naïve-to-memory ratio | 1.02 (0.99, 1.05) | 0.97 (0.85, 1.10) | 1.00 (0.93, 1.08) | 0.97 (0.89, 1.06) |
| CD8+ | 1.00 (0.99, 1.01) | 1.02 (0.99, 1.06) | 1.01 (0.98, 1.05) | 1.01 (0.99, 1.04) |
| CD8+ memory | 1.00 (0.99, 1.02) | 1.03 (1.00, 1.06) | 1.02 (0.99, 1.05) | 1.01 (0.99, 1.03) |
| CD8+ naïve | 0.96 (0.91, 1.01) | 0.85 (0.71, 1.01) | 0.96 (0.86, 1.08) | 0.99 (0.89, 1.11) |
| CD8+ naïve-to-memory ratio | 0.98 (0.87, 1.09) | 0.89 (0.60, 1.09) | 0.97 (0.75, 1.25) | 0.82 (0.61, 1.11) |
| CD4+-to-CD8+ ratio | 0.97 (0.91, 1.03) | 0.99 (0.85, 1.17) | 0.90 (0.75, 1.08) | 0.93 (0.81, 1.06) |
| Treg | 1.06 (1.00, 1.12) | **1.21 (1.05, 1.40)** | 1.04 (0.92, 1.18) | 1.09 (0.96, 1.23) |
| B cell | 0.99 (0.97, 1.02) | 0.98 (0.91, 1.07) | 1.04 (0.98, 1.10) | 0.97 (0.92, 1.03) |
| B cell memory | **1.13 (1.05, 1.21)** | 1.18 (0.97, 1.44) | 0.95 (0.73, 1.23) | **1.18 (1.05, 1.33)** |
| B cell naïve | 0.98 (0.95, 1.01) | 0.96 (0.88, 1.05) | 1.05 (0.99, 1.07) | 0.94 (0.89, 1.00) |
| B cell naïve-to-memory ratio | 0.99 (0.98, 1.00) | 0.97 (0.94, 1.00) | 1.02 (1.00, 1.04) | 0.98 (0.96, 1.00) |
| NLR | 1.00 (0.98, 1.02) | 0.94 (0.86, 1.03) | 0.99 (0.94, 1.04) | 1.01 (0.97, 1.04) |
| Lymphocyte to Monocyte Ratio | 1.00 (1.00, 1.00) | 1.00 (1.00, 1.00) | 1.00 (1.00, 1.00) | 1.03 (0.95, 1.13) |
| White blood cell count (from CBC differential) | 1.02 (0.98, 1.07)  (n = 659) | **1.18 (1.05, 1.32)**  (n = 83) | 1.04 (0.93, 1.17)  (n = 112) | 0.99 (0.90, 1.08)  (n = 171) |

^a^ Excluding hematologic cancers. ^b^ Multivariable models were adjusted for age, sex, BMI, self-reported smoking status, self-reported smoking pack-years, postmenopausal hormone use (creating an ‘NA’ category for males), methylation-derived smoking pack-years, mdNLR (in all models except mdNLR and lymphocyte to monocyte ratio), diabetes status, educational status, and batch effect. ^c^ The breast cancer model also adjusted for self-reported drinking status and did not adjust for sex. ^d^ The prostate cancer model did not adjust for sex or postmenopausal hormone use (men only).

Supplementary Table 2: HRs for Time-Lag Analysis:

HR ^b^ (95% CI) per 1 percent increase in methylation-derived immune cell proportion or 1 unit increase in ratios or white blood cell count

| Methylation-Derived Immune Cell Type or Measure | All Cancer ^b^  (625 cases) | Lung Cancer  (79 cases) | Postmenopausal Breast Cancer ^c^  (106 cases) | Prostate Cancer ^d^  (163 cases) |
| --- | --- | --- | --- | --- |
| CD4+ | 1.00 (0.98, 1.01) | 1.00 (0.96, 1.04) | 1.00 (0.97, 1.03) | 1.00 (0.98, 1.03) |
| CD4+ memory | 0.99 (0.98, 1.01) | 1.00 (0.95, 1.05) | 1.02 (0.98, 1.05) | 1.00 (0.97, 1.03) |
| CD4+ naïve | 1.00 (0.98, 1.02) | 1.00 (0.95, 1.06) | 0.97 (0.93, 1.02) | 1.00 (0.96, 1.05) |
| CD4+ naïve-to-memory ratio | 1.02 (0.98, 1.05) | 0.96 (0.84, 1.09) | 0.99 (0.91, 1.08) | 0.96 (0.88, 1.05) |
| CD8+ | 1.00 (0.99, 1.01) | 1.03 (1.00, 1.07) | 1.01 (0.98, 1.04) | 1.01 (0.99, 1.03) |
| CD8+ memory | 1.00 (0.99, 1.02) | 1.04 (1.00, 1.07) | 1.02 (0.99, 1.05) | 1.01 (0.99, 1.03) |
| CD8+ naïve | 0.95 (0.90, 1.01) | 0.86 (0.72, 1.02) | 0.95 (0.84, 1.07) | 1.00 (0.89, 1.12) |
| CD8+ naïve-to-memory ratio | 0.97 (0.87, 1.09) | 0.89 (0.60, 1.32) | 0.93 (0.70, 1.23) | 0.83 (0.61, 1.14) |
| CD4+-to-CD8+ ratio | 0.98 (0.92, 1.05) | 0.99 (0.85, 1.15) | 0.90 (0.75, 1.09) | 0.94 (0.82, 1.08) |
| Treg | 1.06 (1.00, 1.12) | **1.25 (1.08, 1.44)** | 1.04 (0.91, 1.18) | 1.12 (0.98 (1.26) |
| B cell | 1.00 (0.97, 1.03) | 0.99 (0.91, 1.07) | 1.04 (0.98, 1.11) | 0.98 (0.93, 1.04) |
| B cell memory | **1.14 (1.06, 1.23)** | 1.21 (1.00, 1.48) | 0.95 (0.72, 1.24) | **1.19 (1.05, 1.34)** |
| B cell naïve | 0.98 (0.96, 1.01) | 0.96 (0.88, 1.05) | 1.05 (0.99, 1.11) | 0.95 (0.90, 1.01) |
| B cell naïve-to-memory ratio | 0.99 (0.98, 1.00) | 0.97 (0.94, 1.01) | 1.02 (0.99, 1.04) | 0.98 (0.96, 1.01) |
| NLR | 1.00 (0.98, 1.02) | 0.94 (0.85, 1.03) | 0.99 (0.94, 1.04) | 1.03 (0.99, 1.06) |
| Lymphocyte to Monocyte Ratio | 1.00 (1.00, 1.00) | 1.00 (1.00, 1.00) | 1.00 (1.00, 1.00) | 1.00 (0.88, 1.13) |
| White blood cell count (from CBC differential) | 1.02 (0.98, 1.07)  (n = 616) | **1.20 (1.07, 1.34)**  (n = 78) | 1.04 (0.92, 1.17)  (n = 104) | 0.98 (0.89, 1.07)  (n = 161) |

^a^ Excluding hematologic cancers. ^b^ To assess the possibility of reverse causation, individuals with less than two years of follow-up were excluded from this analysis, regardless of whether they were a case of cancer or lost to follow-up within two years. Multivariable models were adjusted for age, sex, BMI, self-reported smoking status, self-reported smoking pack-years, postmenopausal hormone use (creating an ‘NA’ category for males), methylation-derived smoking pack-years, mdNLR (in all models except mdNLR and lymphocyte to monocyte ratio), and batch effect. ^c^ The breast cancer model also adjusted for self-reported drinking status and did not adjust for sex. ^d^ The prostate cancer model did not adjust for sex or postmenopausal hormone use (men only).

Supplementary Table 3: HRs for Quartile Analysis in All Cancers ^a^:

HR ^a^ (95% CI) for each quartile of methylation-derived immune cell proportion or 1 unit increase in ratios or white blood cell count

|  | Quartile | | | |  | |
| --- | --- | --- | --- | --- | --- | --- |
| Methylation-Derived Immune Cell  Type or Measure | Quartile 1 HR  (95% CI) | Quartile 2 HR  (95% CI) | Quartile 3 HR  (95% CI) | Quartile 4 HR  (95% CI) | | p-trend |
| CD4+ | Ref.  (n = 182) | 0.88 (0.70, 1.10)  (n = 165) | 0.89 (0.71, 1.12)  (n = 168) | 0.89 (0.70, 1.13)  (n = 153) | | 0.33 |
| CD4+ memory | Ref.  (n = 175) | 0.93 (0.74, 1.16)  (n = 161) | 0.90 (0.71, 1.13)  (n = 173) | 0.87 (0.68, 1.11)  (n = 159) | | 0.26 |
| CD4+ naïve | Ref.  (n = 181) | 0.92 (0.74, 1.14)  (n = 166) | 0.96 (0.77, 1.20)  (n = 167) | 0.98 (0.78, 1.24)  (n = 154) | | 0.99 |
| CD4+ naïve-to-memory ratio | Ref.  (n = 168) | 1.15 (0.93, 1.42)  (n = 173) | 0.96 (0.76, 1.21)  (n = 152) | 1.21 (0.96, 1.52)  (n = 175) | | 0.30 |
| CD8+ | Ref.  (n = 180) | 1.01 (0.81, 1.26)  (n = 161) | 0.92 (0.73, 1.15)  (n = 154) | 1.05 (0.83, 1.31)  (n = 173) | | 0.83 |
| CD8+ memory | Ref.  (n = 178) | 0.97 (0.78, 1.21)  (n = 166) | 0.83 (0.66, 1.05)  (n = 146) | 1.05 (0.84, 1.31)  (n = 178) | | 0.49 |
| CD8+ naïve | Ref.  (n = 322) | 1.02 (0.83, 1.25)  (n = 134) | 0.80 (0.64, 1.01)  (n = 105) | 0.83 (0.66, 1.04)  (n = 107) | | 0.09 |
| CD8+ naïve-to-memory ratio | Ref.  (n = 182) | 0.95 (0.77, 1.17)  (n = 172) | 0.83 (0.67, 1.03)  (n = 153) | 0.87 (0.69, 1.09)  (n = 161) | | 0.67 |
| CD4+-to-CD8+ ratio | Ref.  (n = 196) | **0.74 (0.60, 0.91)**  (n = 150) | **0.76 (0.61, 0.94)**  (n = 154) | 0.85 (0.69, 1.06)  (n = 168) | | 0.39 |
| Treg | Ref.  (n = 201) | **1.30 (1.03, 1.64)**  (n = 114) | 1.24 (1.00, 1.53)  (n = 166) | **1.31 (1.04, 1.65)**  (n = 187) | | 0.05 |
| B cell | Ref.  (n = 177) | 0.83 (0.66, 1.04)  (n = 156) | 0.89 (0.71, 1.11)  (n = 170) | 0.93 (0.74, 1.18)  (n = 165) | | 0.61 |
| B cell memory | Ref.  (n = 496) | 0.73 (0.53, 1.01)  (n = 40) | 1.16 (0.88, 1.53)  (n = 60) | **1.47 (1.13, 1.91)**  (n = 72) | | 0.001 |
| B cell naïve | Ref.  (n = 187) | **0.79 (0.63, 0.98)**  (n = 165) | **0.77 (0.61, 0.96)**  (n = 161) | **0.79 (0.62, 0.99)**  (n = 155) | | 0.14 |
| B cell naïve-to-memory ratio | Ref.  (n = 186) | 0.91 (0.74, 1.12)  (n = 181) | **0.76 (0.61, 0.96)**  (n = 150) | 0.81 (0.64, 1.02)  (n = 151) | | 0.08 |
| NLR | Ref.  (n = 154) | 1.00 (0.80, 1.25)  (n = 159) | 1.08 (0.87, 1.34)  (n = 177) | 1.14 (0.91, 1.42)  (n = 178) | | 0.96 |
| Lymphocyte to Monocyte Ratio | Ref.  (n = 177) | 0.90 (0.73, 1.12)  (n = 161) | 1.09 (0.88, 1.35)  (n = 174) | 1.01 (0.80, 1.26)  (n = 156) | | 0.81 |
| White blood cell count (from CBC  differential) | Ref.  (n = 165) | 1.04 (0.83, 1.31)  (n = 149) | 1.16 (0.94, 1.45)  (n = 174) | 1.15 (0.92, 1.44)  (n = 171) | | 0.21 |

^a^ Excluding hematologic cancers. Multivariable models were adjusted for age, sex, BMI, self-reported smoking status, self-reported smoking pack-years, methylation-derived smoking pack-years, postmenopausal hormone use, mdNLR, and batch effect.

Supplementary Table 4: HRs for Tertile Analysis in Lung Cancer:

HR (95% CI) for each tertile of methylation-derived immune cell proportion or 1 unit increase in ratios or white blood cell count

|  | Tertile | | |  |
| --- | --- | --- | --- | --- |
| Methylation-Derived Immune Cell  Type or Measure | Tertile 1 HR  (95% CI) | Tertile 2 HR  (95% CI) | Tertile 3 HR  (95% CI) | p-trend |
| CD4+ | Ref.  (n = 22) | 1.60 (0.90, 2.85)  (n = 38) | 1.10 (0.57, 2.14)  (n = 24) | 0.81 |
| CD4+ memory | Ref.  (n = 22) | 1.42 (0.80, 2.52)  (n = 38) | 0.83 (0.42, 1.63)  (n = 24) | 0.79 |
| CD4+ naïve | Ref.  (n = 34) | 0.70 (0.41, 1.20)  (n = 24) | 0.97 (0.56, 1.69)  (n = 26) | 0.98 |
| CD4+ naïve-to-memory ratio | Ref.  (n = 35) | 0.76 (0.44, 1.31)  (n = 24) | 1.10 (0.62, 1.93)  (n = 25) | 0.56 |
| CD8+ | Ref.  (n = 31) | 0.84 (0.48, 1.47)  (n = 25) | 1.03 (0.59, 1.79)  (n = 28) | 0.16 |
| CD8+ memory | Ref.  (n = 26) | 1.28 (0.74, 2.22)  (n = 31) | 1.27 (0.71, 2.27)  (n = 27) | 0.06 |
| CD8+ naïve | Ref.  (n = 46) | 0.67 (0.40, 1.13)  (n = 23) | **0.47 (0.25, 0.87)**  (n = 15) | 0.06 |
| CD8+ naïve-to-memory ratio | Ref.  (n = 34) | 0.87 (0.52, 1.46)  (n = 28) | 0.57 (0.32, 1.01)  (n = 22) | 0.49 |
| CD4+-to-CD8+ ratio | Ref.  (n = 35) | **0.54 (0.31, 0.94)**  (n = 20) | 0.73 (0.44, 1.22)  (n = 29) | 0.82 |
| Treg | Ref.  (n = 20) | 1.26 (0.69, 2.28)  (n = 26) | 1.42 (0.77, 2.63)  (n = 38) | 0.01 |
| B cell | Ref.  (n = 27) | 0.83 (0.47, 1.46)  (n = 27) | 0.99 (0.56, 1.77)  (n = 30) | 0.66 |
| B cell memory | Ref.  (n = 54) | 1.27 (0.68, 2.36)  (n = 14) | 1.56 (0.84, 2.89)  (n = 16) | 0.60 |
| B cell naïve | Ref.  (n = 30) | 0.81 (0.47, 1.39)  (n = 29) | 0.74 (0.41, 1.32)  (n = 25) | 0.29 |
| B cell naïve-to-memory ratio | Ref.  (n = 37) | 0.77 (0.46, 1.30)  (n = 27) | **0.53 (0.29, 0.97)**  (n = 20) | 0.06 |
| NLR | Ref.  (n = 19) | **1.78 (1.01, 3.14)**  (n = 35) | 1.17 (0.64, 2.12)  (n = 30) | 0.21 |
| Lymphocyte to Monocyte Ratio | Ref.  (n = 26) | 1.53 (0.91, 2.57)  (n = 34) | 1.29 (0.72, 2.33)  (n = 24) | 0.99 |
| White blood cell count (from CBC  differential) | Ref.  (n = 15) | 1.73 (0.91, 3.27)  (n = 28) | **2.67 (1.43, 4.97)**  (n = 40) | 0.002 |

Multivariable models were adjusted for age, sex, BMI, self-reported smoking status, self-reported smoking pack-years, methylation-derived smoking pack-years, postmenopausal hormone use, mdNLR, and batch effect.

Supplementary Table 5: HRs for Tertile Analysis in Postmenopausal Breast Cancer:

HR (95% CI) for each tertile of methylation-derived immune cell proportion or 1 unit increase in ratios or white blood cell count

|  | Tertile | | |  |
| --- | --- | --- | --- | --- |
| Methylation-Derived Immune Cell  Type or Measure | Tertile 1 HR  (95% CI) | Tertile 2 HR  (95% CI) | Tertile 3 HR  (95% CI) | p-trend |
| CD4+ | Ref.  (n = 33) | 0.97 (0.59, 1.58)  (n = 40) | 0.85 (0.51, 1.41)  (n = 41) | 0.78 |
| CD4+ memory | Ref.  (n = 37) | 0.84 (0.50, 1.39)  (n = 32) | 1.08 (0.65, 1.79)  (n = 45) | 0.41 |
| CD4+ naïve | Ref.  (n = 33) | 1.02 (0.64, 1.63)  (n = 44) | 0.72 (0.44, 1.20)  (n = 37) | 0.18 |
| CD4+ naïve-to-memory ratio | Ref.  (n = 41) | 0.71 (0.45, 1.15)  (n = 34) | 0.76 (0.47, 1.23)  (n = 39) | 0.95 |
| CD8+ | Ref.  (n = 41) | 0.66 (0.40, 1.07)  (n = 30) | 0.93 (0.59, 1.47)  (n = 43) | 0.34 |
| CD8+ memory | Ref.  (n = 42) | **0.61 (0.37, 0.99)**  (n = 27) | 1.10 (0.71, 1.72)  (n = 45) | 0.21 |
| CD8+ naïve | Ref.  (n = 62) | **0.36 (0.20, 0.63)**  (n = 16) | **0.64 (0.41, 0.99)**  (n = 36) | 0.47 |
| CD8+ naïve-to-memory ratio | Ref.  (n = 45) | **0.60 (0.37, 0.96)**  (n = 28) | 0.73 (0.46, 1.15)  (n = 41) | 0.76 |
| CD4+-to-CD8+ ratio | Ref.  (n = 34) | 1.09 (0.70, 1.72)  (n = 46) | 0.83 (0.51, 1.36)  (n = 34) | 0.23 |
| Treg | Ref.  (n = 34) | 0.75 (0.44, 1.28)  (n = 25) | 1.49 (0.92, 2.43)  (n = 55) | 0.51 |
| B cell | Ref.  (n = 29) | 0.85 (0.51, 1.43)  (n = 34) | 1.14 (0.70, 1.86)  (n = 51) | 0.22 |
| B cell memory | Ref.  (n = 94) | 0.44 (0.19, 1.01)  (n = 6) | 1.20 (0.65, 2.19)  (n = 14) | 0.72 |
| B cell naïve | Ref.  (n = 25) | 1.05 (0.61, 1.78)  (n = 36) | 1.40 (0.84, 2.34)  (n = 53) | 0.17 |
| B cell naïve-to-memory ratio | Ref.  (n = 30) | 0.82 (0.49, 1.36)  (n = 33) | 1.12 (0.68, 1.84)  (n = 51) | 0.18 |
| NLR | Ref.  (n = 42) | 0.97 (0.62, 1.51)  (n = 38) | 1.02 (0.65, 1.61)  (n = 34) | 0.70 |
| Lymphocyte to Monocyte Ratio | Ref.  (n = 27) | 0.96 (0.57, 1.61)  (n = 33) | 1.31 (0.81, 2.09)  (n = 54) | 0.84 |
| White blood cell count (from CBC  differential) | Ref.  (n = 36) | 1.10 (0.70, 1.75)  (n = 40) | 1.22 (0.75, 1.98)  (n = 36) | 0.37 |

Multivariable models were adjusted for BMI, self-reported smoking status, self-reported smoking pack-years, methylation-derived smoking pack-years, self-reported drinking status, postmenopausal hormone use, mdNLR, and batch effect.

Supplementary Table 6: HRs for Tertile Analysis in Prostate Cancer:

HR (95% CI) for each tertile of methylation-derived immune cell proportion or 1 unit increase in ratios or white blood cell count

|  | Tertile | | |  |
| --- | --- | --- | --- | --- |
| Methylation-Derived Immune Cell  Type or Measure | Tertile 1 HR  (95% CI) | Tertile 2 HR  (95% CI) | Tertile 3 HR  (95% CI) | p-trend |
| CD4+ | Ref.  (n = 68) | 1.08 (0.74, 1.57)  (n = 59) | 1.10 (0.74, 1.65)  (n = 46) | 0.69 |
| CD4+ memory | Ref.  (n = 59) | 1.19 (0.81, 1.75)  (n = 62) | 1.13 (0.74, 1.72)  (n = 52) | 0.95 |
| CD4+ naïve | Ref.  (n = 75) | 0.85 (0.59, 1.22)  (n = 53) | 1.02 (0.70, 1.51)  (n = 45) | 0.56 |
| CD4+ naïve-to-memory ratio | Ref.  (n = 64) | 1.13 (0.79, 1.64)  (n = 60) | 0.95 (0.64, 1.41)  (n = 49) | 0.61 |
| CD8+ | Ref.  (n = 56) | 1.25 (0.84, 1.87)  (n = 53) | 1.40 (0.95, 2.06)  (n = 64) | 0.31 |
| CD8+ memory | Ref.  (n = 57) | 1.06 (0.71, 1.59)  (n = 49) | 1.24 (0.85, 1.82)  (n = 67) | 0.38 |
| CD8+ naïve | Ref.  (n = 84) | **1.47 (1.05, 2.07)**  (n = 60) | 0.89 (0.57, 1.38)  (n = 29) | 0.98 |
| CD8+ naïve-to-memory ratio | Ref.  (n = 63) | 1.21 (0.86, 1.72)  (n = 67) | 0.79 (0.52, 1.20)  (n = 43) | 0.24 |
| CD4+-to-CD8+ ratio | Ref.  (n = 71) | 0.87 (0.60, 1.24)  (n = 53) | 0.84 (0.58, 1.22)  (n = 49) | 0.35 |
| Treg | Ref.  (n = 55) | **1.59 (1.10, 2.31)**  (n = 66) | 1.34 (0.87, 2.06)  (n = 52) | 0.20 |
| B cell | Ref.  (n = 81) | 0.83 (0.57, 1.20)  (n = 50) | 0.87 (0.59, 1.29)  (n = 42) | 0.32 |
| B cell memory | Ref.  (n = 127) | 0.76 (0.45, 1.30)  (n = 16) | 1.27 (0.83, 1.95)  (n = 30) | 0.01 |
| B cell naïve | Ref.  (n = 85) | 0.77 (0.54, 1.11)  (n = 51) | 0.70 (0.47, 1.06)  (n = 37) | 0.06 |
| B cell naïve-to-memory ratio | Ref.  (n = 85) | **0.68 (0.47, 0.98)**  (n = 50) | 0.72 (0.48, 1.08)  (n = 38) | 0.10 |
| NLR | Ref.  (n = 55) | 0.96 (0.66, 1.40)  (n = 56) | 0.91 (0.63, 1.31)  (n = 62) | 0.59 |
| Lymphocyte to Monocyte Ratio | Ref.  (n = 71) | 1.12 (0.79, 1.59)  (n 60) | 1.16 (0.79, 1.71)  (n = 42) | 0.47 |
| White blood cell count (from CBC  differential) | Ref.  (n = 66) | 1.12 (0.78, 1.63)  (n = 56) | 0.95 (0.64, 1.40)  (n = 49) | 0.80 |

Multivariable models were adjusted for BMI, self-reported smoking status, self-reported smoking pack-years, methylation-derived smoking pack-years, mdNLR, and batch effect.

Supplementary Table 7: HRs for Sensitivity Analysis Including Total Leukocyte Count in All Cancers ^a^ and Lung Cancer:

HR ^b^ (95% CI) per 1 percent increase in methylation-derived immune cell proportion or 1 unit increase in ratios or white blood cell count

|  | Cancer Type | | | |
| --- | --- | --- | --- | --- |
|  | All Cancers | | Lung Cancer | |
| Methylation-Derived Immune Cell Type or Measure | With WBC in Model  (659 cases) | Without WBC in Model  (668 cases) | With WBC in Model  (83 cases) | Without WBC in Model  (84 cases) |
| CD4+ naïve-to-memory ratio | 1.02 (0.98, 1.05) | 1.02 (0.99, 1.05) | 0.95 (0.83, 1.09) | 0.96 (0.85, 1.09) |
| CD8+ naïve | 0.96 (0.91, 1.01) | 0.95 (0.90, 1.01) | 0.85 (0.71, 1.01) | 0.85 (0.71, 1.01) |
| CD8+ naïve-to-memory ratio | 0.98 (0.88, 1.09) | 0.97 (0.87, 1.09) | 0.85 (0.57, 1.26) | 0.87 (0.59, 1.29) |
| CD4+-to-CD8+ ratio | 0.97 (0.91, 1.04) | 0.98 (0.92, 1.05) | 0.96 (0.82, 1.13) | 0.98 (0.84, 1.15) |
| Treg | 1.06 (1.00, 1.12) | 1.06 (1.00, 1.12) | **1.22 (1.06, 1.41)** | **1.22 (1.06, 1.41)** |
| B cell memory | **1.13 (1.05, 1.22)** | **1.13 (1.05, 1.22)** | **1.24 (1.02, 1.50)** | 1.20 (0.99, 1.46) |

^a^ Excluding hematologic cancers. This model performed a sensitivity analysis to assess whether white blood cell count was independent of other relevant immune cell type proportions and measures. ^b^ Multivariable models were adjusted for sex, BMI, self-reported smoking status, self-reported smoking pack-years, methylation-derived smoking pack-years, postmenopausal hormone use, mdNLR, white blood cell count, and batch effect.

Supplementary Table 8: HRs for Sex-Stratified Model in All Cancers ^a^:

HR ^b^ (95% CI) per 1 percent increase in methylation-derived immune cell proportion or 1 unit increase in ratios or white blood cell count

|  | Sex | |  |
| --- | --- | --- | --- |
| Methylation-Derived Immune Cell  Type or Measure | Females  (343 cases) | Males  (325 cases) | p-value for interaction |
| CD4+ | 0.99 (0.97, 1.00) | 1.00 (0.98, 1.02) | 0.25 |
| CD4+ memory | 0.99 (0.97, 1.01) | 0.99 (0.97, 1.02) | 0.48 |
| CD4+ naïve | 0.99 (0.97, 1.01) | 1.01 (0.98, 1.04) | 0.31 |
| CD4+ naïve-to-memory ratio | 1.03 (0.99, 1.07) | 0.98 (0.92, 1.05) | 0.13 |
| CD8+ | 0.99 (0.97, 1.01) | 1.01 (0.99, 1.02) | 0.12 |
| CD8+ memory | 1.00 (0.98, 1.02) | 1.01 (0.99, 1.02) | 0.19 |
| CD8+ naïve | 0.96 (0.90, 1.02) | 0.95 (0.87, 1.04) | 0.63 |
| CD8+ naïve-to-memory ratio | 0.98 (0.85, 1.14) | 0.97 (0.82, 1.16) | 0.61 |
| CD4+-to-CD8+ ratio | 0.98 (0.90, 1.07) | 0.97 (0.88, 1.05) | 0.71 |
| Treg | 1.06 (0.99, 1.14) | 1.03 (0.95, 1.13) | 0.08 |
| B cell | 1.01 (0.97, 1.04) | 0.97 (0.93, 1.01) | 0.22 |
| B cell memory | 1.09 (0.97, 1.22) | **1.16 (1.05, 1.28)** | 0.22 |
| B cell naïve | 1.00 (0.96, 1.03) | **0.95 (0.91, 0.99)** | 0.05 |
| B cell naïve-to-memory ratio | 1.00 (0.98, 1.01) | 0.98 (0.97, 1.00) | 0.08 |
| NLR | 1.00 (0.97, 1.02) | 1.00 (0.97, 1.03) | 0.67 |
| Lymphocyte to Monocyte Ratio | 1.00 (1.00, 1.00) | 1.02 (0.96, 1.10) | 0.29 |
| White blood cell count (from CBC  differential) | **1.09 (1.02, 1.16)**  (n = 337) | 0.98 (0.92, 1.05)  (n = 322) | 0.01 |

^a^ Excluding hematologic cancers. ^b^ Multivariable models were adjusted for BMI, age, self-reported smoking status, self-reported smoking pack-years, methylation-derived smoking pack-years, mdNLR, post-menopausal hormone status (for women only), and batch effect.

Supplementary Table 9: HRs for Sex-Stratified Model in Lung Cancer:

HR (95% CI) per 1 percent increase in methylation-derived immune cell proportion or 1 unit increase in ratios or white blood cell count

|  | Sex | |  |
| --- | --- | --- | --- |
| Methylation-Derived Immune Cell Type or Measure | Females  (37 cases) | Males  (47 cases) | p-value for  interaction |
| CD4+ | 0.99 (0.93, 1.04) | 1.01 (0.96, 1.06) | 0.22 |
| CD4+ memory | 0.99 (0.93, 1.06) | 1.01 (0.95, 1.07) | 0.22 |
| CD4+ naïve | 0.98 (0.91, 1.06) | 1.02 (0.94, 1.11) | 0.67 |
| CD4+ naïve-to-memory ratio | 0.90 (0.75, 1.08) | 1.01 (0.82, 1.24) | 0.85 |
| CD8+ | 0.99 (0.93, 1.06) | 1.04 (1.00, 1.08) | 0.16 |
| CD8+ memory | 1.01 (0.95, 1.07) | 1.04 (1.00, 1.08) | 0.21 |
| CD8+ naïve | 0.88 (0.71, 1.09) | 0.77 (0.57, 1.06) | 0.26 |
| CD8+ naïve-to-memory ratio | 0.84 (0.49, 1.42) | 0.91 (0.52, 1.61) | 0.74 |
| CD4+-to-CD8+ ratio | 0.99 (0.77, 1.26) | 0.99 (0.81, 1.21) | 0.99 |
| Treg | 1.13 (0.93, 1.37) | 1.16 (0.92, 1.45) | 0.01 |
| B cell | 0.93 (0.82, 1.05) | 1.02 (0.91, 1.13) | 0.92 |
| B cell memory | 1.10 (0.78, 1.53) | **1.34 (1.04, 1.71)** | 0.19 |
| B cell naïve | 0.92 (0.81, 1.05) | 0.97 (0.84, 1.09) | 0.59 |
| B cell naïve-to-memory ratio | 0.95 (0.91, 1.00) | 0.97 (0.93, 1.02) | 0.50 |
| NLR | 0.91 (0.77, 1.07) | 0.98 (0.89, 1.09) | 0.76 |
| Lymphocyte to Monocyte Ratio | 1.00 (1.00, 1.00) | 1.08 (0.91, 1.29) | 0.19 |
| White blood cell count (from CBC differential) | **1.31 (1.09, 1.58)**  (n = 36) | 1.09 (0.94, 1.26)  (n = 47) | 0.12 |

Multivariable models were adjusted for BMI, age, self-reported smoking status, self-reported smoking pack-years, methylation-derived smoking pack-years, mdNLR, post-menopausal hormone status (for women only), and batch effect.

Supplementary Figure 1: All Cancer Spline Plots:

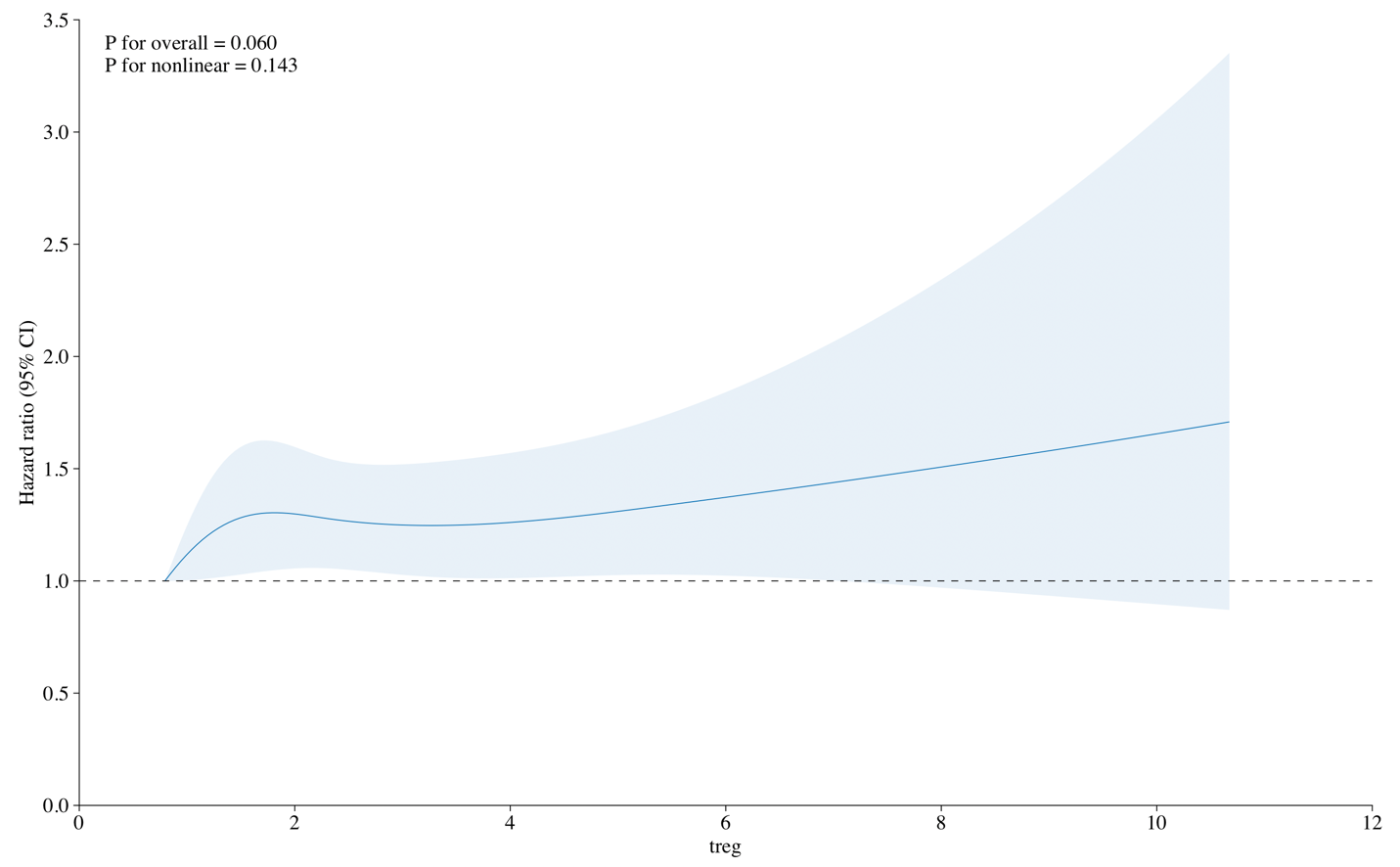

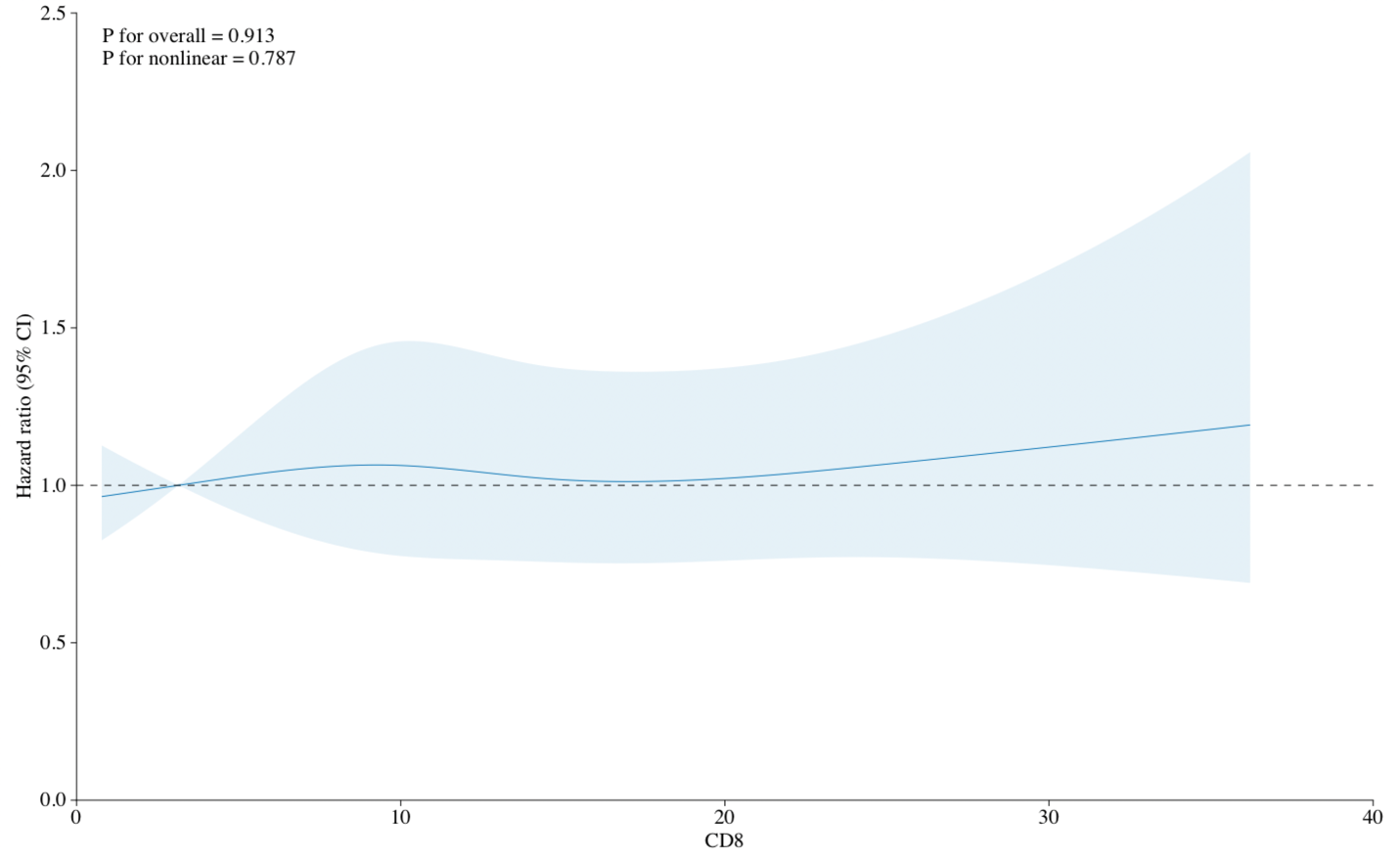

These spline plots assess the changes in the hazard ratios at various proportions of Tregs and CD8+ cells in all cancers. The P for nonlinear for Tregs of 0.143 and P for nonlinear of 0.787 for CD8+ cells indicates that a linear model is likely an appropriate means of representing these associations.

Supplementary Figure 2: Lung Cancer Spline Plots:

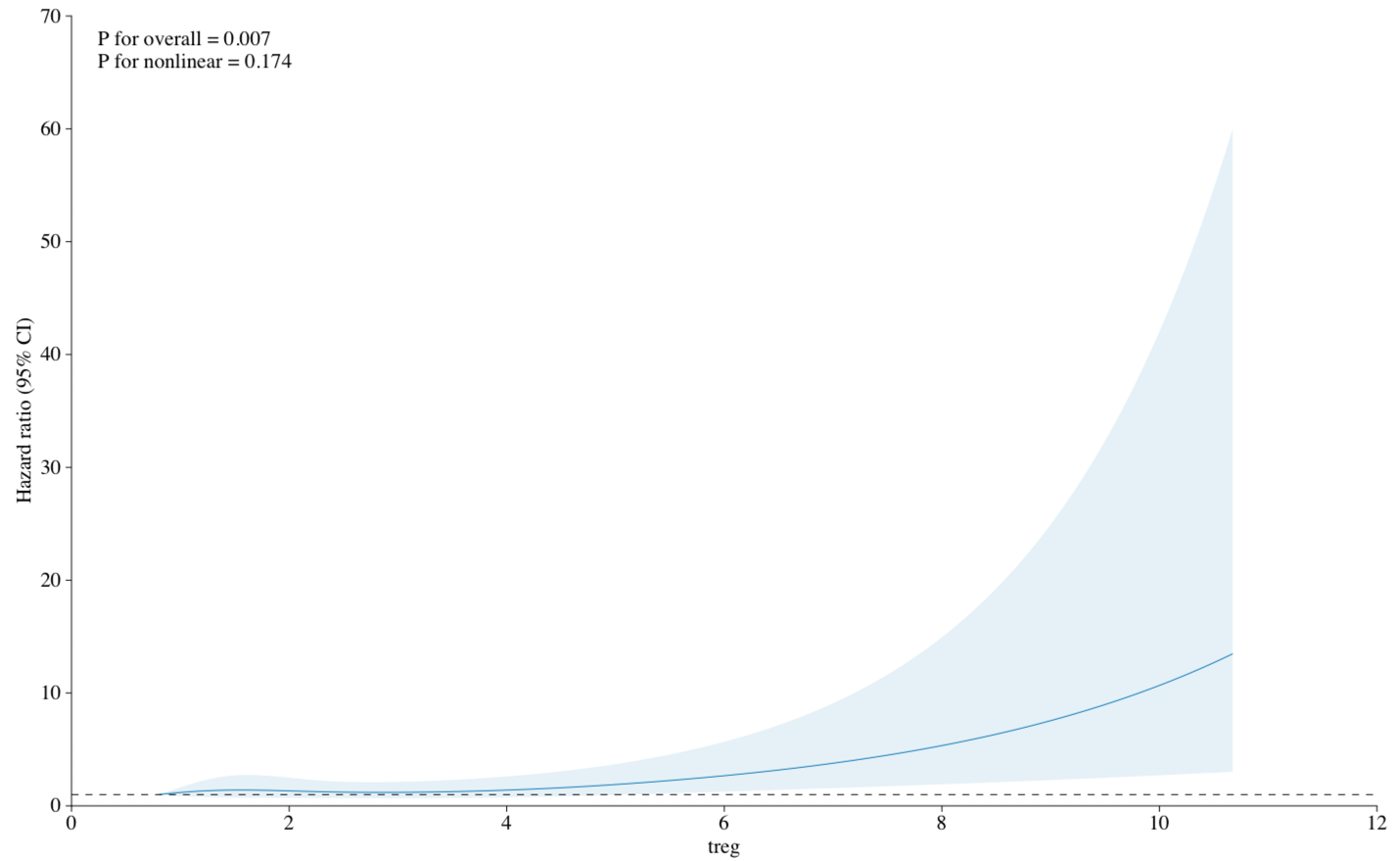

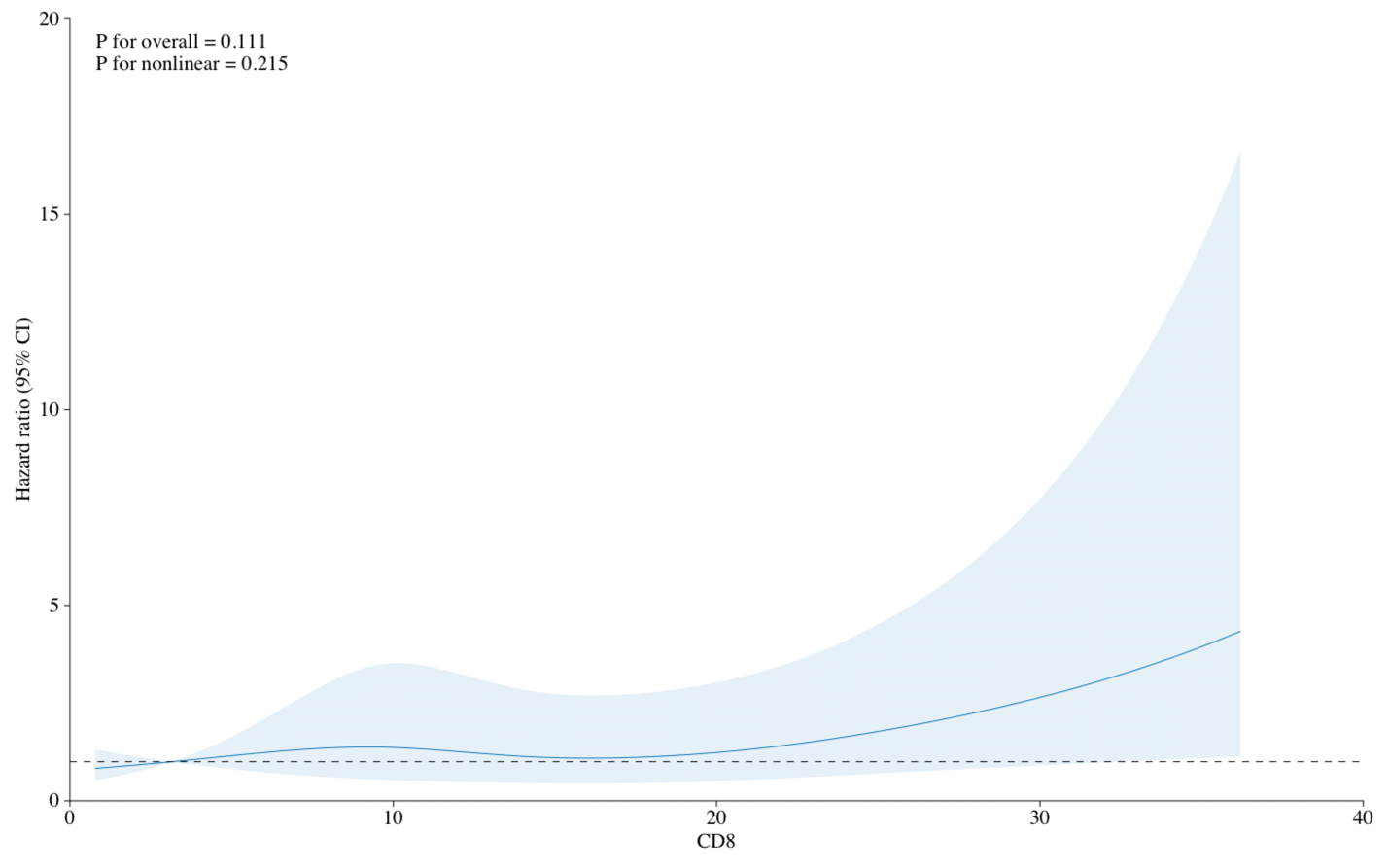

These spline plots assess the changes in the hazard ratios at various proportions of Tregs and CD8+ cells in lung cancer. The P for nonlinear for Tregs of 0.174 and P for nonlinear of 0.215 for CD8+ cells indicates that a linear model is likely an appropriate means of representing these associations.

Supplementary Figure 3: Postmenopausal Breast Cancer Spline Plots:

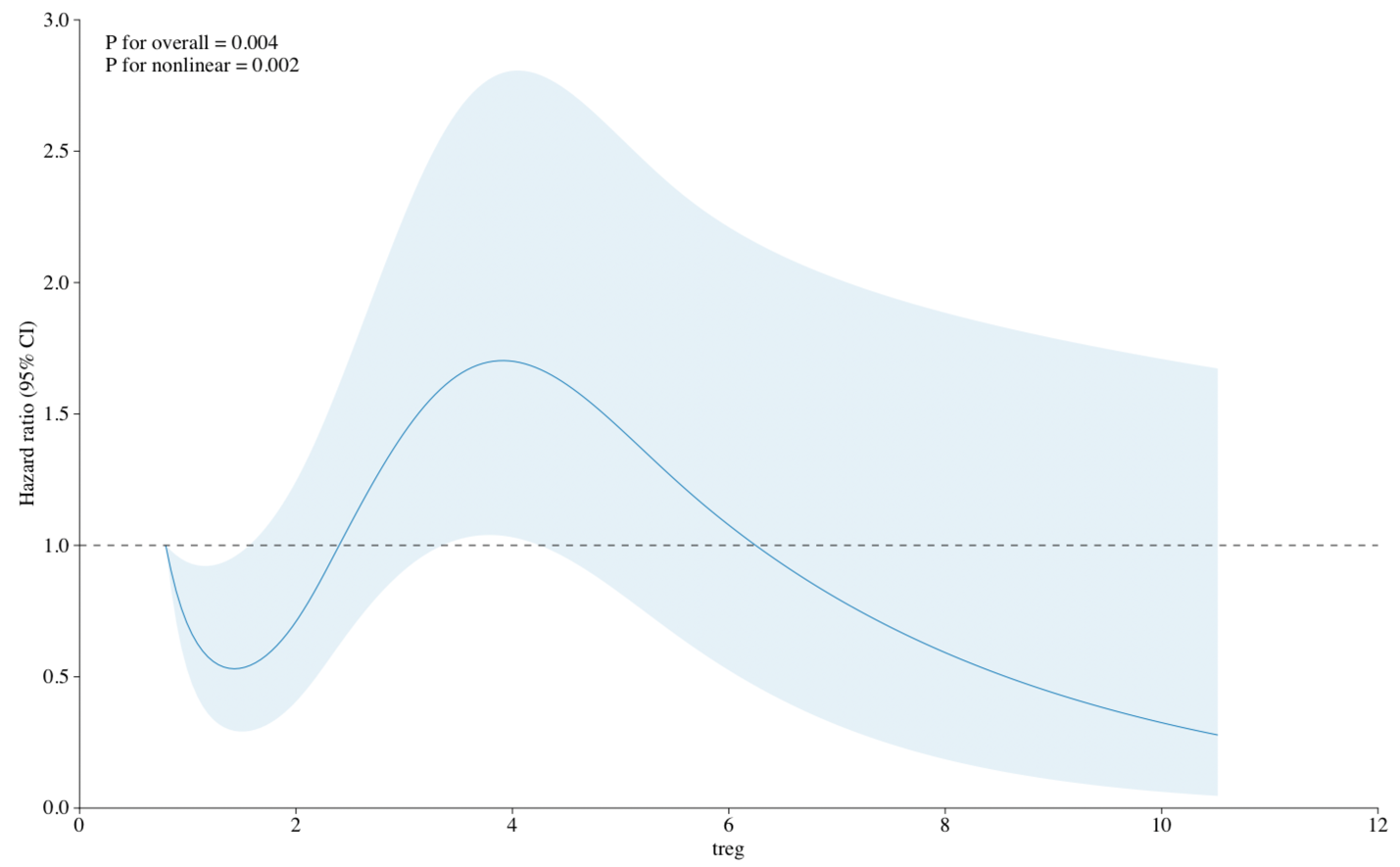

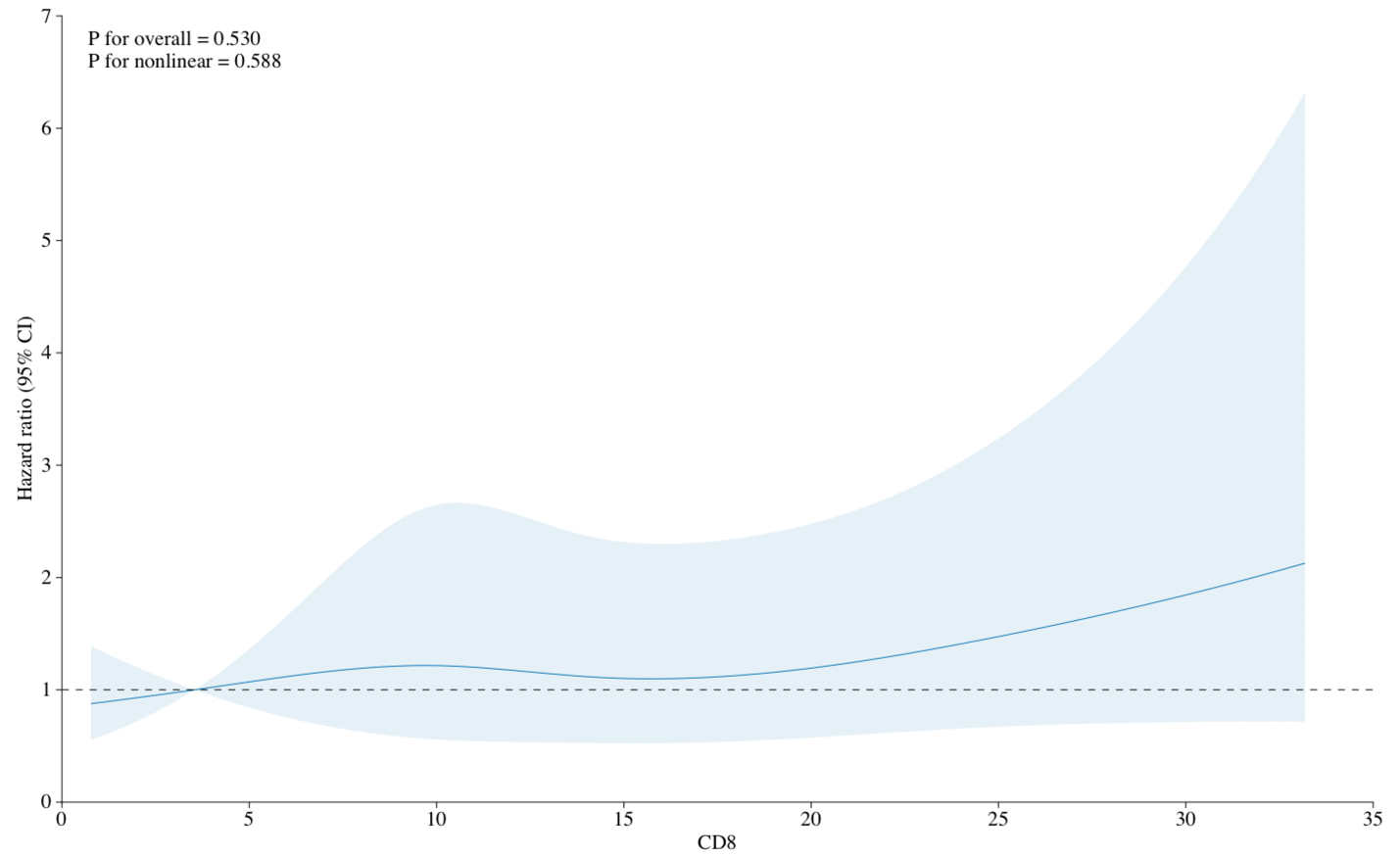

These spline plots assess the changes in the hazard ratios at various proportions of Tregs and CD8+ cells in breast cancer. The P for nonlinear for Tregs of 0.002 and P for nonlinear of 0.588 for CD8+ cells call into question the appropriateness of a linear model for assessing this particular relationship.

Supplementary Figure 4: Prostate Cancer Spline Plots:

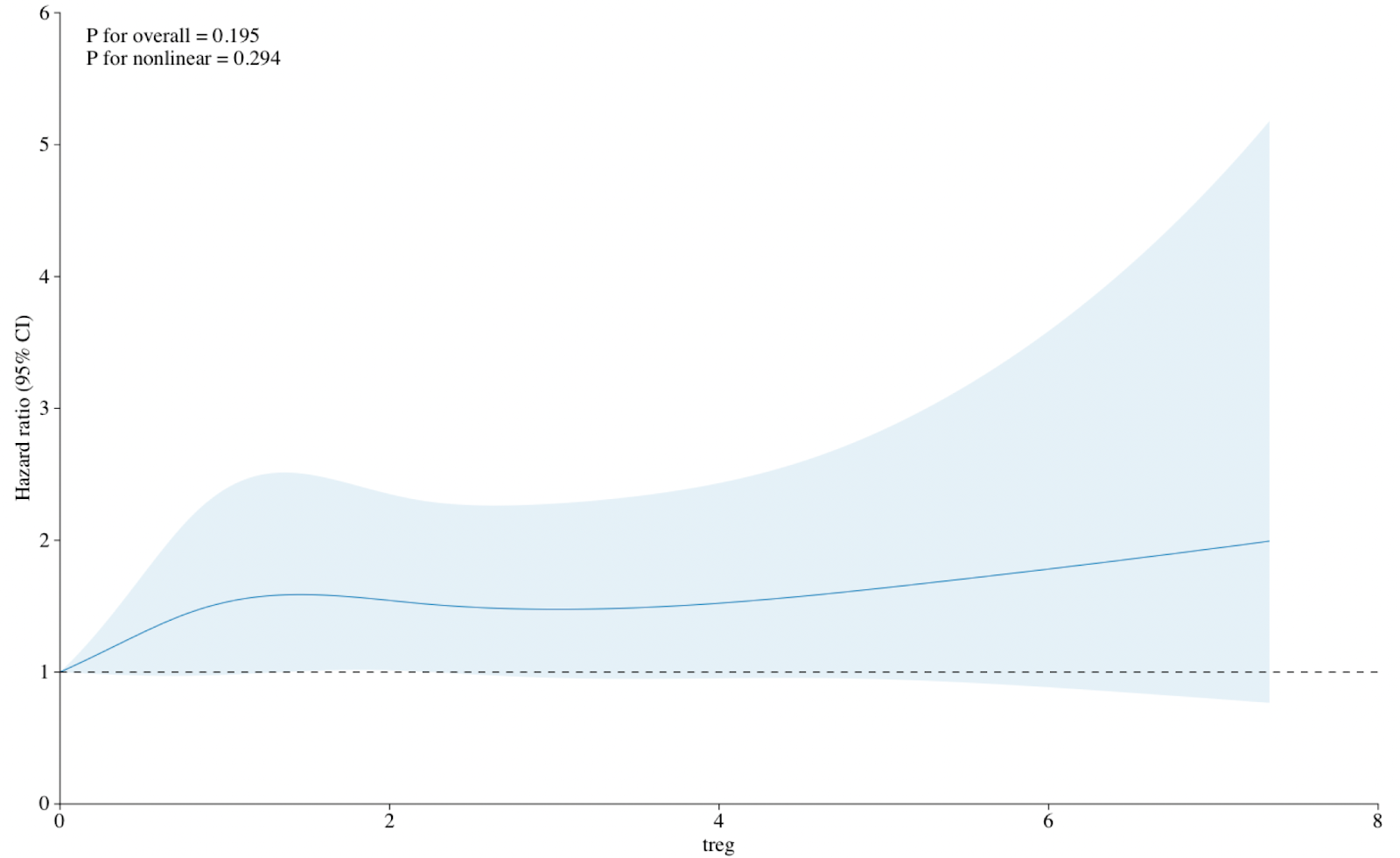

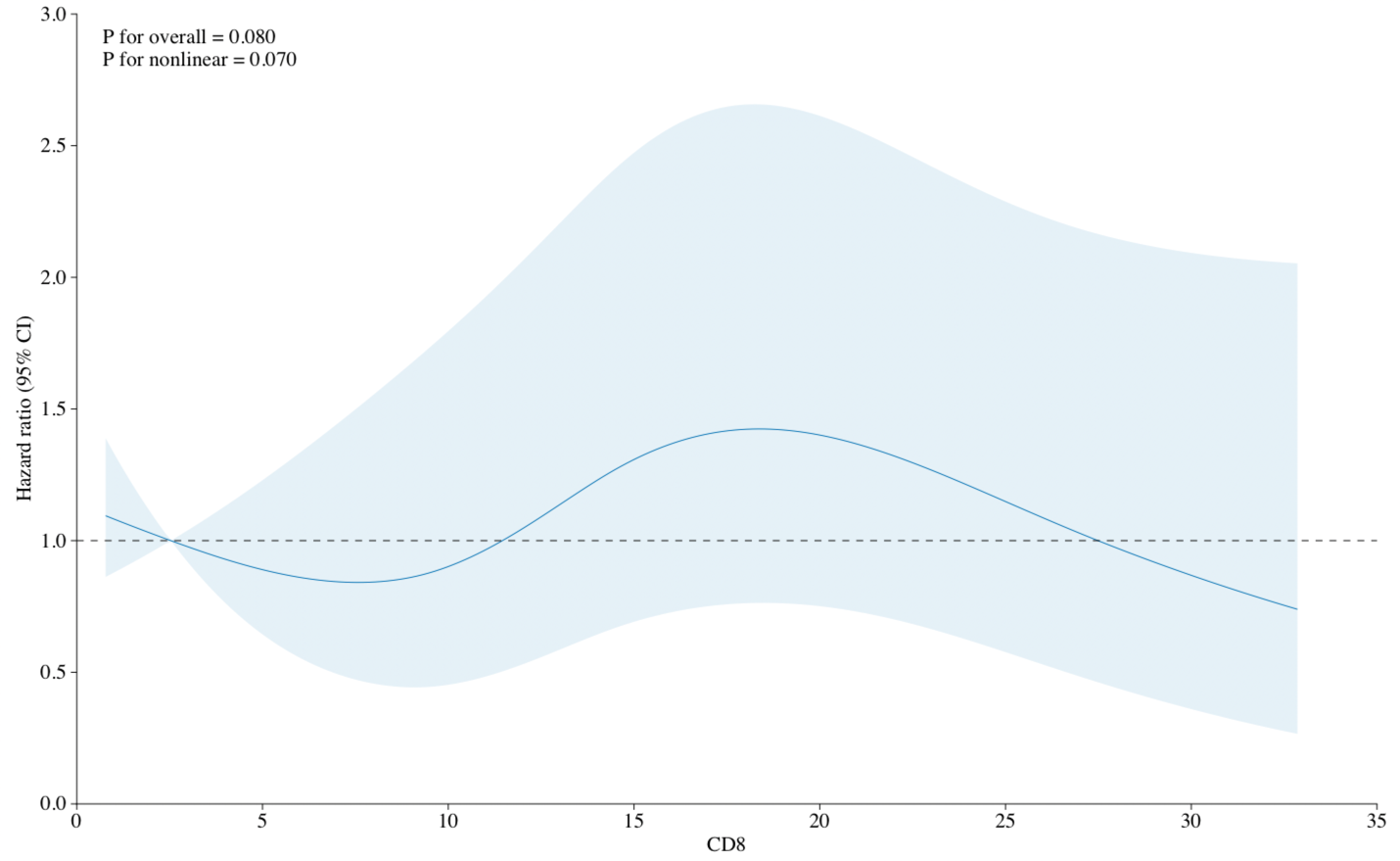

These spline plots assess the changes in the hazard ratios at various proportions of Tregs and CD8+ cells in prostate cancer. The P for nonlinear for Tregs of 0.294 and P for nonlinear of 0.070 for CD8+ cells indicates that a linear model is likely an appropriate means of representing these associations.
